# Serum proteome analysis of systemic JIA and related pulmonary alveolar proteinosis identifies distinct inflammatory programs and biomarkers

**DOI:** 10.1101/2021.01.20.21250141

**Authors:** Guangbo Chen, Gail Deutsch, Grant Schulert, Hong Zheng, SoRi Jang, Bruce Trapnell, Pui Lee, Claudia Macaubas, Katherine Ho, Corinne Schneider, Vivian E. Saper, Adriana Almeida de Jesus, Mark Krasnow, Alexei Grom, Raphaela Goldbach-Mansky, Purvesh Khatri, Elizabeth D Mellins, Scott W. Canna

**Affiliations:** Institute for Immunity, Transplantation and Infection, School of Medicine, Stanford University, Stanford, CA, USA; Pathology, Seattle Children’s Hospital and University of Washington Medical Center, Seattle, WA, USA; Cincinnati Children’s Hospital Medical Center, University of Cincinnati College of Medicine, Cincinnati, OH, USA; Center for Biomedical Informatics Research, Department of Medicine, School of Medicine, Stanford University, Stanford, CA, USA; Biochemistry, Howard Hughes Medical Institute, Stanford University School of Medicine, Stanford, CA, USA; Pediatric Rheumatology, Boston Children’s Hospital and Harvard School of Medicine, Boston, MA, USA; Pediatrics, Program in Immunology, School of Medicine, Stanford University, Stanford, CA, USA; Pediatrics, UPMC Children’s Hospital & University of Pittsburgh Medical Center, Pittsburgh, PA, USA; Pediatrics, School of Medicine, Stanford University, Stanford, California, USA; Translational Autoinflammatory Disease Section, National Institute of Allergy and Infectious Diseases, Bethesda, MD, USA; Pediatric Rheumatology, The Children’s Hospital of Philadelphia, Philadelphia, PA, USA

**Keywords:** interstitial lung disease, systemic JIA, macrophage activation syndrome, ICAM5

## Abstract

**Objectives:** Recent observations in systemic Juvenile Idiopathic Arthritis (sJIA) suggest an increasing incidence of high-mortality interstitial lung disease, characterized by a variant of pulmonary alveolar proteinosis (PAP). Co-occurrence of macrophage activation syndrome (MAS) and PAP in sJIA suggested a shared pathology, but sJIA-PAP patients also commonly experience features of drug reaction such as atypical rashes and eosinophilia. We sought to investigate immunopathology and identify biomarkers in sJIA, MAS, and sJIA-PAP.

**Methods:** We used SOMAscan to measure >1300 analytes in sera from healthy controls and patients with sJIA, MAS, sJIA-PAP and other related diseases. We verified selected findings by ELISA and lung immunostaining. Because the proteome of a sample may reflect multiple states (sJIA, MAS, sJIA-PAP), we used regression modeling to identify subsets of altered proteins associated with each state. We tested key findings in a validation cohort.

**Results:** Proteome alterations in active sJIA and MAS overlapped substantially, including known sJIA biomarkers like SAA and S100A9, and novel elevations of heat shock proteins and glycolytic enzymes. IL-18 was elevated in all sJIA groups, particularly MAS and sJIA-PAP. We also identified an MAS-independent sJIA-PAP signature notable for elevated ICAM5, MMP7, and allergic/eosinophilic chemokines, which were all previously associated with lung damage. Immunohistochemistry localized ICAM5 and MMP7 in sJIA-PAP lung. ICAM5’s ability to distinguish sJIA-PAP from sJIA/MAS was independently validated.

**Conclusions:** Serum proteins support an sJIA-to-MAS continuum, help distinguish sJIA, sJIA/MAS, and sJIA-PAP, and suggest etiologic hypotheses. Select biomarkers, such as ICAM5, could aid in early detection and management of sJIA-PAP.

## Introduction

Systemic juvenile idiopathic arthritis (sJIA) is a chronic inflammatory disease of childhood characterized by a combination of systemic inflammation, quotidian fever, evanescent rash, adenopathy/organomegly, serositis, and arthritis[1]. Its adult equivalent is adult-onset Still’s disease (AOSD)[2]. Macrophage activation syndrome (MAS) is a life-threatening form of secondary hemophagocytic lymphohistiocytosis (HLH) that complicates about 10% of patients with sJIA[3]. It is characterized by cytokine storm, very high serum ferritin, progression to organ failure, and a mortality of up to 17%. Active sJIA and MAS may share a common etiology, representing a spectrum of disease severity[4].

For decades, pleuritis and pleural effusions were the typical lung manifestations identified in sJIA [1, 5]. In recent years, pediatric rheumatologists have increasingly observed sJIA patients who developed acute digital clubbing and an insidious parenchymal lung disease[6, 7]. The predominant pathology of this new lung disease is a variant of pulmonary alveolar proteinosis (PAP)/endogenous lipoid pneumonia (ELP), with increased inflammatory infiltrate, pulmonary arterial wall thickening, but little fibrosis[7]. Nearly all cases with this pathology were exposed to IL-1 and/or IL-6 inhibition, and the drug-exposed group demonstrated other clinical features atypical for sJIA, such as acute erythematous clubbing, atypical rash, eosinophilia, and anaphylaxis to tocilizumab[7]. Though the initial descriptions were associated with severe sJIA (with MAS in 80%)[6], larger follow-up series suggested that MAS at sJIA onset was not associated with the unusual clinical features of the sJIA-PAP group, and some patients with treatment-responsive sJIA nevertheless developed lung disease. However, 78% had overt or subclinical MAS at the detection of or during lung disease, suggesting lung disease may stimulate MAS[7].

To better understand the underlying pathology and relationships between sJIA and its complications MAS and sJIA-PAP, we assembled serum proteomes from a multi-center cohort of sJIA patients with or without these complications and relevant comparators (monogenic autoinflammatory diseases and PAP from other causes). We also sought to identify biomarkers that might aid in detecting or monitoring PAP in sJIA.

## Methods

### Cohort & Disease Definitions

Several names have been used for the syndrome of digital clubbing and PAP-variant parenchymal lung disease observed in sJIA[6-9]. Herein, we use the term “sJIA-PAP” to identify patients known or strongly suspected to have this constellation of features **(Supplementary Table 1)**. Clinical data and serum from sJIA-PAP, healthy controls, inactive sJIA, active sJIA, MAS, hereditary/autoimmune PAP, STING-associated Vasculopathy of Infancy (SAVI, all with interstitial lung disease, prior to Jak inhibitor therapy), Neonatal-Onset Multisystem Inflammatory Disease (NOMID, prior to IL-1 blockade), and NLRC4-MAS were collected under ongoing protocols using established diagnostic, classification, or genetic criteria. Notably, arthritis was not required for classification as sJIA. The study included a discovery cohort comprised of all groups above, as well as a validation cohort consisting of only samples from controls and sJIA patients (see **Table 1, Supplemental Table 1**, and **Supplemental Methods)**.

**Table 1:** Group Definitions and Demographics of the Discovery Cohort

| Groups* | Clinical Definition | Patient N | Sample N** | Age, median (IQR) | Female % |
| --- | --- | --- | --- | --- | --- |
| Healthy Ctrl | No known pulmonary or rheumatic/autoinflammatory disease | 21 | 21 | 8.9 (1.4-17) | 37% (7/19) \$ |
| Inactive sJIA | Inactive at time of sampling per submitting investigator | 28 | 30 | 12 (7-16) | 64%(18/28) |
| Active sJIA | Active per submitting investigator but did not meet MAS criteria at time of sampling | 24 | 25 | 12 (8-14) | 58%(14/24) |
| MAS | Met MAS criteria at time of sampling | 10 | 12 | 9.4 (4.1-16) | 70%(7/10) |
| sJIA-PAP | Had radiologic evidence of lung disease. See Table S1 For details | 10\$\$ | 30 | 5.2 (4.3-8.1) | 60%(6/10) |
| subgroup: sJIA-PAP FCLow | Ferritin AND CRP elevated (thresholds in Figure S1) | 8 | 18 | 4.4 (3.8-6.4) | 63%(5/8) |
| subgroup: sJIA-PAP FCHigh | Either ferritin OR CRP not elevated | 6 | 12 | 6.5 (5-8.6) | 50%(3/6) |
| PAP | PAP due to anti-GM-CSF autoantibodies (10) or genetic causes (4) without sJIA or known rheumatic/autoinflammatory disease | 14 | 14 | 34 (17-47) | 57%(8/14) |
| SAVI | STING-associated Vasculopathy with onset in Infancy. All patients had radiologic ILD, prior to treatment with jak inhibitors | 4 | 4 | 15 (10-18) | 100%(4/4) |
| NOMID | Neonatal Onset Multi-System Inflammatory Disease, prior to treatment with IL-1 inhibitors | 5 | 5 | 9.2 (8.6-18) | 80%(4/5) |
| Inact NLRC4 MAS | Inactive per submitting investigator | 2 | 5 | 5.3 (3.2-7.4) | 100%(2/2) |
| NLRC4 MAS | Active MAS per submitting investigator | 2 | 5 | 4.8 (2.5-7.2) | 100%(2/2) |
| Total |  | 110 | 151 |  |  |

\*: The bold font highlights the main patient groups subjected to LIMMA analysis
\*\* : When multiple samples were taken from the same patient in the same patient group, the SOMAscan data were merged by averaging.
\$: Two healthy controls were missing data on sex.
Four patients overlapped between the FCLow and FCHigh subgroups.
All sJIA patients met modified ILAR criteria [DeWitt et al., Arth Care Res, 2012]

### Serum protein analysis

For the discovery cohort, we profiled serum samples by SOMAscan assay (SOMAlogic, Boulder, CO), an aptamer-based proteomics platform[10], in collaboration with the NIH Center for Human Immunology. 1271 analytes were evaluated, with most being mapped to a single protein (exceptions are listed in **Supplementary Tables 2-4**). IL-18 was assessed by Luminex. CXCL9 and IL-18 binding protein (IL-18BP) were measured by both SOMAscan and Luminex to verify reproducibility (**Supplementary Figure 1**). We performed ICAM-5 ELISA on remnant sera from the discovery cohort as a technical verification. Serum and plasma samples from an independent validation cohort were assayed for IL-18 and CXCL9 by Luminex and ICAM-5 and MMP7 by ELISA. **Table 1, Supplementary Table 1**, and **Supplemental Methods** contain further details.

### Linear Regression-Based Modeling Analysis

A schematic overview of the study is shown in **Supplementary Figure 2A**. Patients’ disease, and therefore their proteomes, may reflect multiple disease components (e.g., patients with sJIA-PAP may have an active sJIA disease component). Thus, our analysis required distinguishing patient groups from disease components, with some overlap of disease and component names (e.g., the active sJIA group vs. the sJIA disease component). Likewise, components can contribute to the proteomes of multiple disease groups (e.g., MAS disease component in MAS and sJIA-PAP patient samples). To capture the serum proteome alterations attributed to a specific disease component, we used LIMMA (Linear Models for Microarray Data), in which the overall proteome alteration was regressed against the disease component(s) present in each individual patient **(Supplementary Figure 2B-C)**. We used these disease components to assign disease (sJIA, MAS, sJIA/PAP) activity scores to individual samples. For details on LIMMA, disease component construction, and calculating disease activity scores, see **Supplemental Methods**.

## Results

### A discovery cohort including sJIA, MAS, sJIA-PAP and related inflammatory conditions

We included 151 serum samples from 120 patients for proteomic profiling using SOMAscan. The patients represented 10 patient groups, based on clinical characterizations **(Table 1, Supplementary Table 1)**. Most samples and patients were from five main groups that we analyzed by linear regression: healthy controls (n=21), inactive sJIA (28), active sJIA (24), MAS (10), and sJIA-PAP (10), **(Supplemental Methods)**. The sJIA-PAP samples were divided into those with high levels of both ferritin and CRP (sJIA-PAP^FCHi^) and those without (sJIA-PAP^FCLo^, **Supplementary Figures 3A-B)**. All sJIA-PAP patient samples were collected during or following exposure to IL-1 and or IL-6 inhibitors **(Supplementary Table 1)**.

### Determine the proteome changes associated with each disease component

In all patient sera studied, the proteome in a single sample may reflect contributions of multiple concurrent pathologic processes **(Supplementary Figure 2)**. A central goal of this study was to capture the serum proteome alterations attributed to a specific disease component (sJIA, MAS or sJIA-PAP), as distinguished from the disease groups (see above and **Supplemental Methods**). We used Limma to determine the statistically significant differences associated with each disease component **(Figure 1A-C, Supplementary Figures 2B-C)**.

**Figure 1:**
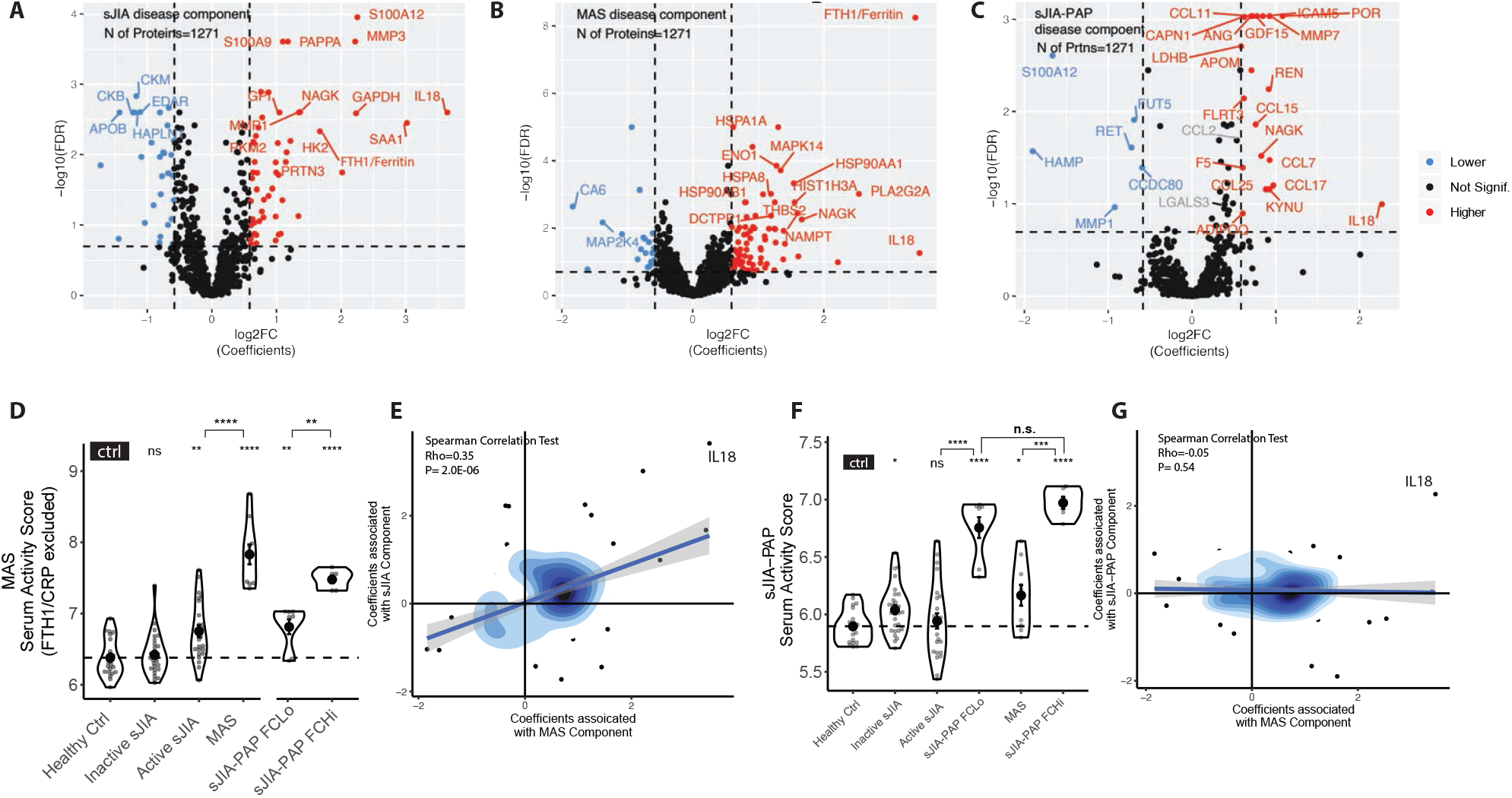
Serum proteome profile associated with active sJIA/MAS/sJIA-PAP. A,B,C) Volcano plots highlight the proteins (shown by gene names) with significantly changed abundance in the sJIA (A) or MAS (B) or sJIA-PAP disease component (C). Significance thresholds are represented by the dashed lines: false discovery rate [FDR (adjusted p-value)] < 20% and fold change > 1.5. D) The MAS serum activity score (calculated without CRP or ferritin, see methods) across different groups is shown. sJIA-PAP is sub-grouped by ferritin and CRP values (see **Table 1**). Dashed horizontal line indicates median value of healthy controls. Between-group comparisons were performed using Wilcoxon signed-rank test without the assumption of normal distribution, and reflect comparisons to the control group unless otherwise indicated. We used the Benjamini-Hochberg procedure to adjust p-values. *, p<0.05; **, p<0.01; ***, p<0.001; ****, p< 0.0001. E) Correlation between the coefficients (Log2FC) assigned by the LIMMA model to the sJIA and MAS disease components (see **Supplemental Methods**) for the 174 proteins identified as significantly altered in at least one of the three disease components (sJIA, MAS, or sJIA-PAP). F and G are similar to D and E, but plot the sJIA-PAP disease component.

### MAS further exaggerates many molecular changes present in sJIA

We first characterized the sJIA and MAS components. For the sJIA component (active sJIA versus healthy controls), we found 86 significantly altered proteins, with most (85%) elevated in the sJIA group **(Figure 1A, Supplementary Table 2)**. Many of these proteins, including ferritin (*FTH1*), IL-18, CRP and S100A12 **(Supplementary Figures 3C-F)**, have been previously associated with sJIA using other techniques, supporting the validity of SOMAscan measurements. We also identified several proteins not typically associated with sJIA, such as MMP3 (a protease induced by IL-6[11]), and various metabolic enzymes, including GAPDH, NAGK, HK, and GPI.

We also compared profiles of inactive sJIA patients to healthy controls. Seven proteins were significantly different **(Supplementary Figure 4)**, including the IL-10 induced monokine CCL16, consistent with a state of compensated inflammation in inactive sJIA[12].

The analysis of the MAS component compared patients with active sJIA to those with MAS and identified 105 significantly different proteins **(Figure 1B, Supplementary Table 3)**. These included many glycolytic enzymes (e.g., GAPDH, LDH) and several chaperone proteins (e.g., HSP70, HSP90). We calculated a MAS serum activity score to quantify the MAS component within each sample (see **Supplemental Methods**). We excluded CRP and ferritin from the score, as these were often used by submitting investigators to define MAS. As expected, the MAS score (even without CRP and ferritin) was significantly greater in MAS than sJIA **(Figure 1D)** and was significantly correlated with ferritin (Spearman correlation test, rho=0.57, p<2.2e-16) and CRP (rho=0.5, p=1.5e-09) **(Supplementary Figures 5A-B)**. The MAS serum activity score was also higher in patients with active versus inactive NLRC4-MAS **(Supplementary Figure 5C)**. In linear regression model, each protein received a coefficient reflecting its contribution to the component **(Supplemental Methods)**. We observed a strong correlation between the protein abundance coefficients associated with sJIA and MAS disease components **(Figure 1E, Supplementary Figure 5D)**, suggesting MAS further exaggerated a serum proteome already altered in sJIA.

### Proteins involved in leukocyte-mediated immunity and cellular metabolism are elevated in the sJIA and MAS serum proteomes

Examining the top altered proteins in the sJIA and MAS disease components **(Supplementary Tables 2-4)**, we found a few proteins whose functional relationship was novel or unclear. Specifically, heat shock proteins Hsp70 (HSPA1A, HSPA8) and Hsp90 (HSP90AA1, HSP90AB1) were upregulated in many patients, particularly in MAS **(Supplementary Table 3, Figure 2A)**. These proteins function as intracellular molecular chaperones, but they may also be extracellular damage-associated molecular patterns (DAMPs)[13]. Additionally, many glycolytic enzymes, like ENO1 and GAPDH, were strongly elevated in MAS and sJIA **(Figures 2B)**. We performed Gene Ontology (GO) term enrichment analysis on the proteins significantly changed for each disease component **(Supplementary Table 5)**. Consistent with systemic immune activation, proteins involved in GO term “leukocyte mediated immunity” **(Figure 2C)** were elevated in both the sJIA and MAS disease components. In addition, we found the sJIA and MAS disease components were enriched (only sJIA component met statistical significance) for proteins involved in GO term “monocarboxylic acid metabolic process” **(Figure 2D, Supplementary Table 5)**.

**Figure 2:**
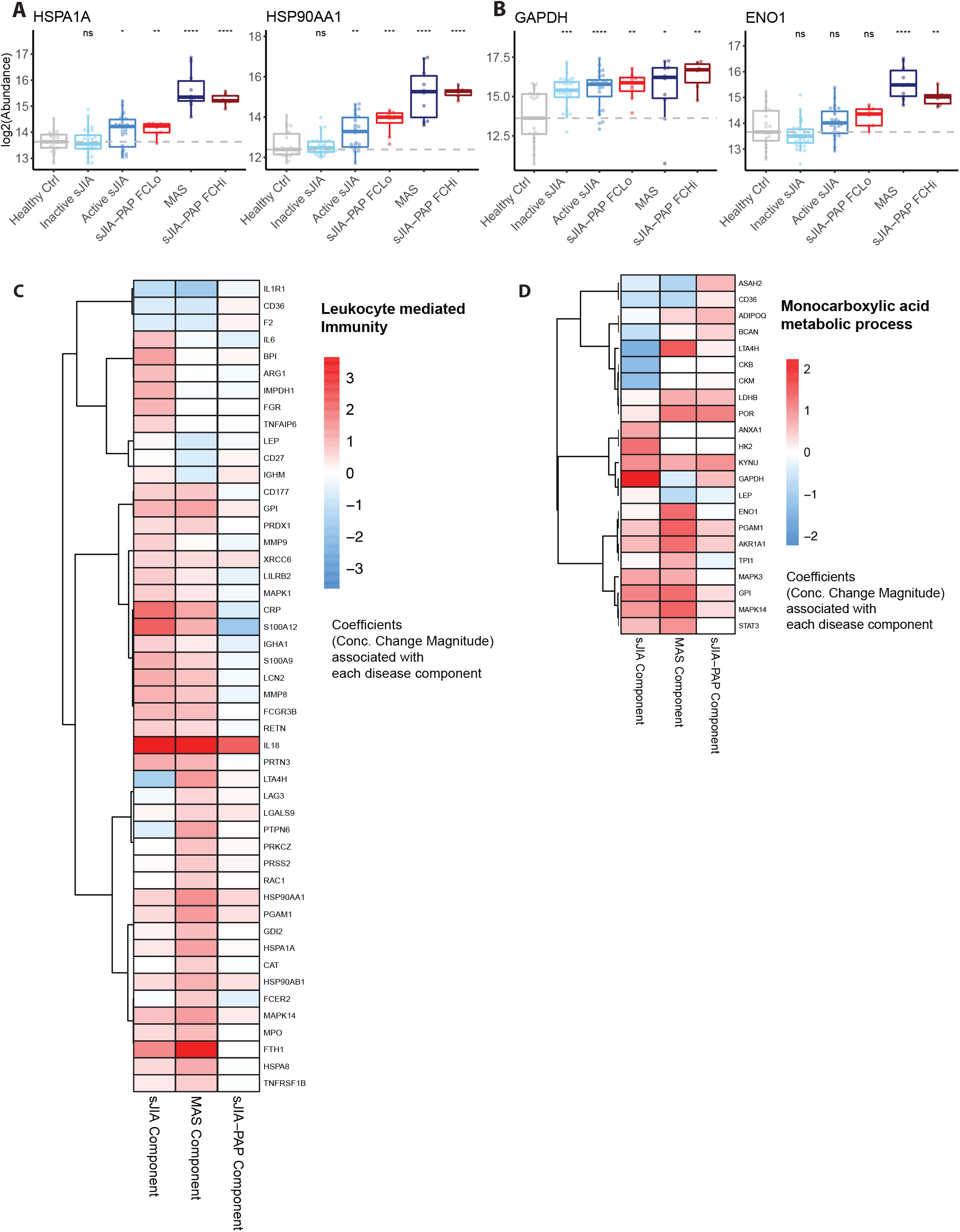
Protein functional groups for different disease components. A) Examples of significantly elevated heat-shock proteins (HSP1A, HSP90AA1) and proteins involved in glycolytic process (GAPDH, ENO1) were plotted for different patient groups. Comparisons between each indicated group and the healthy control group were performed using Wilcoxon signed-rank test with p-values adjusted by Benjamini-Hochberg Procedure. *, p<0.05; **, p<0.01; ***, p<0.001; ****, p< 0.0001. Dashed horizontal line indicates median value of healthy controls. C-D) Proteins present in SOMAscan for two different functional groups defined by Gene Ontology (GO) terms (C: Leukocyte mediated immunity, D: Monocarboxylic acid metabolic process. GO term enrichment results are provided in **Supplementary Table 5**). The heatmap shows the coefficients of each protein assigned by the LIMMA model to each disease component, with disease components and genes clustered by Euclidean distances. To facilitate analysis, gene names are presented but represent protein targets, see **Supplemental Methods**.

### Serum alterations associated with lung disease can occur independent of MAS activity in patients with sJIA-PAP

Active sJIA or MAS did not appear to sufficiently explain sJIA-PAP disease activity, as we observed a low MAS score in many sJIA-PAP samples, and previous clinical data showed sJIA-PAP could arise in the absence of MAS [7]. Therefore, we sought to identify the proteins driving an sJIA-PAP disease component. To track whether sJIA-PAP samples with strong MAS features biased or confounded identification of the sJIA-PAP component, the sJIA-PAP samples were divided into those with high levels of both ferritin and CRP (sJIA-PAP^FCHi^) and those without (sJIA-PAP^FCLo^). The active sJIA and sJIA-PAP^FCLo^ groups had comparable MAS serum activity scores. The sJIA-PAP^FCHi^ group had significantly higher MAS serum activity scores than the active sJIA or sJIA-PAP^FCLo^ groups but trended toward lower MAS serum activity scores than the MAS group **(Figure 1D)**. This suggests significant smoldering MAS activity in some sJIA-PAP samples even though none were obtained during periods meeting clinical MAS criteria. Comparing sJIA-PAP patients to sJIA and MAS patients identified 26 proteins (20 up, 6 down) significantly associated with the sJIA-PAP disease component, including ICAM5, MMP7, and CCL11/Eotaxin-1 (**Figure 1C**, see **Supplemental Methods** on adjustment for MAS activity confounding). Using these proteins, we calculated the sJIA-PAP serum activity score (see **Supplemental Methods**) and found it was higher in sJIA-PAP samples than in either active sJIA or MAS, as expected **(Figure 1F)**. Unlike the MAS serum activity score, the sJIA-PAP serum activity score did not differ between the sJIA-PAP^FCLo^ and sJIA-PAP^FCHi^ groups, and both groups had substantially higher sJIA-PAP serum activity than the MAS group **(Figure 1F, Supplementary Figure 6A)**. The small elevation of the sJIA-PAP activity score in the MAS group was entirely attributable to IL-18 **(Supplementary Figure 6A vs. B)**. As expected by the model design, there was no correlation between the protein abundance coefficients of the sJIA-PAP-component and those of the MAS component **(Figure 1G, Supplementary Figure 6C)**. Notably, the sJIA-PAP serum activity score was higher in sJIA-PAP than in primary PAP caused by GM-CSF-neutralizing autoantibodies or surfactant-related mutations **(Supplementary Figures 6A-B)**.

Half of the sJIA-PAP patients had four or more timepoints analyzed by SOMAscan. When we traced the disease component-specific serum activity scores longitudinally, the sJIA-PAP serum activity score did not correlate with sJIA or MAS serum activity scores **(Supplementary Figure 7)**. Overall, these findings corroborate clinical observations suggesting that sJIA-PAP may be triggered by a mechanism distinct from sJIA, MAS, and other well-characterized PAP conditions.

### sJIA-PAP serum proteome profile is characterized by elevations in IL-18 and type II chemokines

Different types of inflammation are characterized by distinct serum cytokine and chemokine profiles. The changes in cytokine and chemokine levels in serum proteome profiles associated with active sJIA and MAS components appeared similar **(Figure 3A)**, in line with our prior proteome-wide analysis. Only IL-18 significantly contributed to all three components, confirming its association with sJIA/MAS and suggesting an independent role in sJIA-PAP **(Supplementary Figure 8)**.

**Figure 3:**
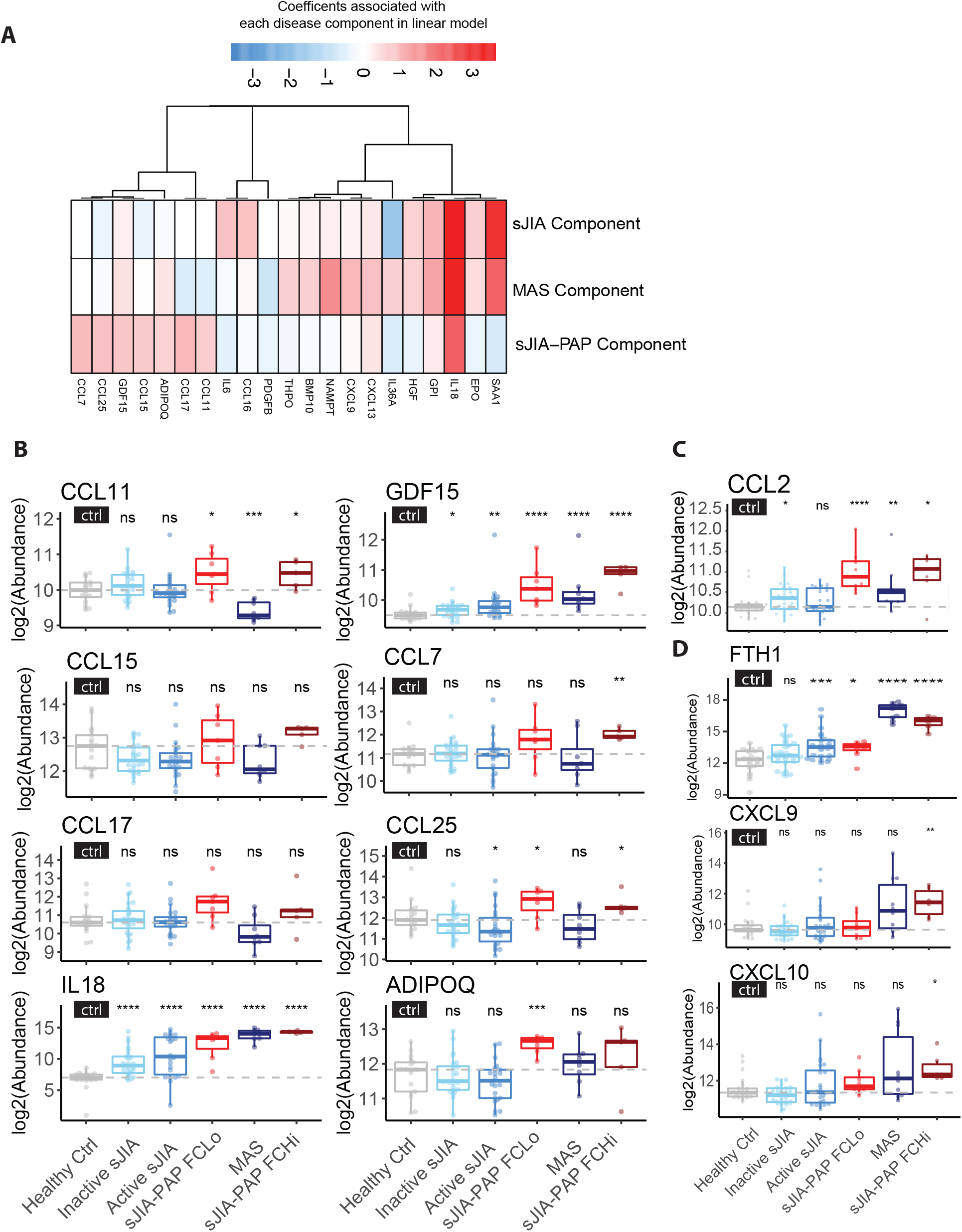
Cytokine/chemokine serum abundance differs between sJIA-PAP and sJIA/MAS. A) The heatmap presents the coefficients of concentration change from the LIMMA analysis for cytokines/chemokines associated with each disease component in the linear regression model. Cytokines/chemokines shown are those that reached significance in at least one disease component and also were associated with the respective GO term (see **Supplemental Methods**). B) Eight significantly-altered cytokines/chemokines in the sJIA-PAP disease component were plotted by different patient groups (see **Supplementary Figure 9** for groups beyond the main disease groups). Comparisons between each indicated group and the healthy control group were performed using Wilcoxon signed-rank test with p value adjusted by Benjamini-Hochberg Procedure. *, p<0.05; **, p<0.01; ***, p<0.001; ****, p< 0.0001. C) CCL2, which approaches significance for sJIA-PAP disease component is also shown. D) CXCL9 and CXCL10, two interferon-inducible chemokines are shown. Dashed horizontal lines indicate median values of healthy controls. To facilitate analysis, gene names are presented but represent protein targets, see **Supplemental Methods**.

Among the cytokines/chemokines associated with the sJIA-PAP disease activity component, most strongly associated were CCL11 (eotaxin-1) and CCL17 (TARC), which did not contribute significantly to the sJIA or MAS components **(Figure 3B, Supplementary Figure 9)**. CCL17/TARC, CCL11/eotaxin-1, and CCL2/MCP-1 are potent chemoattractants for Th2 cells, eosinophils, and myeloid cells, respectively. They were similarly increased in both the sJIA-PAP^FCHi^ and sJIA-PAP^FCLo^ groups, although CCL2 (a chemokine associated with lung disease[14]) did not meet the pre-defined significance threshold in Limma analysis **(Figure 3B-C, Supplementary Figure 9, Supplementary Table 4)**. The MAS-associated, interferon-inducible chemokines CXCL9 and 10 showed similar trends as ferritin and were not routinely elevated in sJIA-PAP **(Figure 3D)**. Several chemokines that contribute to the sJIA-PAP activity score (including CCL11 and CCL17) were not elevated in autoimmune/hereditary PAP **(Supplementary Figures 6A and 9)**, suggesting different etiologies.

### ICAM5 is a potential biomarker for sJIA-PAP

ICAM5 was one of the proteins most significantly associated with sJIA-PAP in the discovery cohort **(Figure 1C, Supplementary Figure 10)**. To verify the SOMAscan ICAM5 findings, we measured ICAM5 by ELISA in 80 remnant samples previously profiled with SOMAscan. As with CXCL9 and IL-18BP **(Supplementary Figure 1)**, we found a strong correlation between ELISA and SOMAscan results **(Supplementary Figure 11)**. Accordingly, only sJIA-PAP samples showed ICAM5 elevation by ELISA, although ELISA showed poorer sensitivity than SOMAscan at low concentrations. ICAM5 function has been best-characterized in the nervous system due to high expression in the brain[15]. However, an autopsy dataset of healthy adults (Genome-Tissue Expression)[16] showed ICAM5 expression in the lung comparable to that seen in the CNS **(Figure 4A)**. MMP7, a matrix metallopeptidase (MMP) that efficiently cleaves ICAM5 *in vitro*[17], was also significantly elevated in sJIA-PAP sera **(Figure 1C, Supplementary Figure 10)**, and we found a significant correlation between serum concentrations of ICAM5 and MMP7 **(Figure 4B)**. In the SOMAscan cohort, ICAM5 serum concentration alone was capable of distinguishing sJIA-PAP from inactive sJIA, active sJIA or MAS cases **(Figure 4C)**.

**Figure 4:**
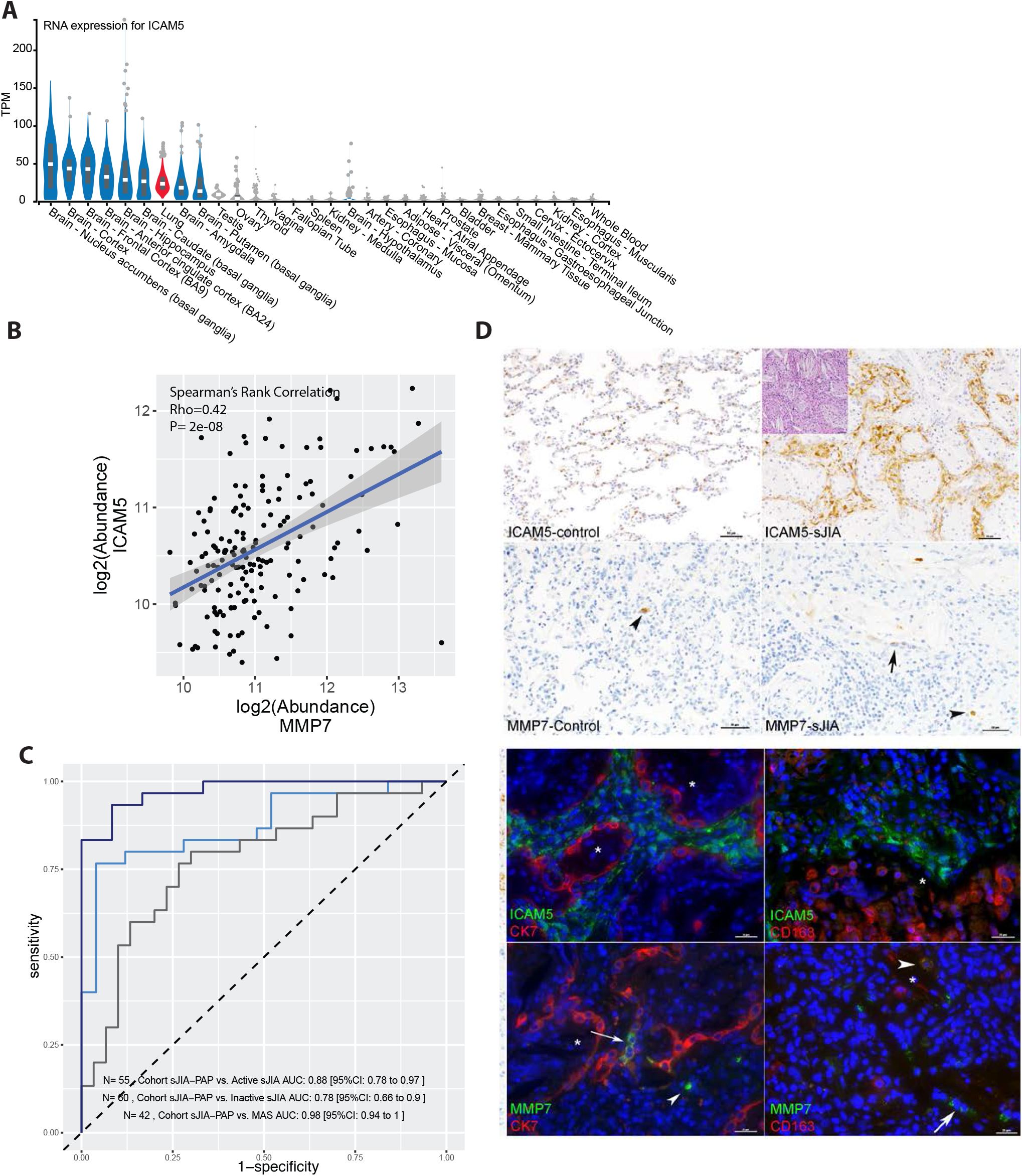
Pulmonary localization of ICAM5 and MMP7, and their performance in the discovery cohort. A) The 30 organs with highest-expression of *ICAM5* mRNA from an autopsy cohort (Genotype-Tissue Expression, GTEx); expression in lung samples is depicted in red. B) The correlation of MMP7 and ICAM5 protein levels among all sJIA serum samples. C) ROC curves using ICAM5 to classify different comparisons in the discovery cohort. D) Staining of ICAM5 and MMP7 in control and sJIA lung. Inset shows characteristic PAP/ELP histology in an sJIA-PAP patient. ICAM5 protein expression in lung interstitial fibroblast cells in contrast with epithelial cells (marked by cytokeratin, CK7) and macrophages (CD163). MMP7 is expressed in control and sJIA-PAP lung by the indicated epithelial cells (arrows) and macrophages and hematopoietic cells (arrowheads). * denotes alveolar lumen. Nuclei counterstained with DAPI (blue). All immunofluorescence images are from sJIA-PAP. See also **Supplementary Figures 12** and **13**.

To determine whether proteins identified by SOMAscan profiling of serum samples were also expressed in lung tissue, we used immunostaining to evaluate lung biopsies from 8 patients with sJIA-PAP, 3 with genetic disorders in surfactant metabolism (*SFTPC, NKX2*.*1*, and *GATA2*), 1 with idiopathic ILD, and 3 controls (asthma/aspiration, bronchopneumonia, and normal lung adjacent to tumor). Consistent with our prior evaluation of sJIA-PAP histology[7], lungs from sJIA-PAP patients exhibited the characteristic pattern of pulmonary alveolar proteinosis (PAP) and/or endogenous lipoid pneumonia (ELP)[7] with mixed acute and chronic inflammatory cells and rare interstitial fibrosis **(Supplementary Table 6 & Figure 4D)**. Lung pathology from the patients with genetic disorders in surfactant metabolism showed a similar spectrum of PAP/ELP with more prominent fibrosis.

All samples showed membrane-associated ICAM5 that was largely restricted to interstitial fibroblasts, confirmed by double-labeling with the epithelial marker cytokeratin 7 **(Figure 4D, Supplementary Figure 12)**. ICAM5 expression was especially prominent in Case 1 (sJIA-PAP) and Case 9 (GATA2) (Supplementary Table 6), and overall proportional to the number of interstitial fibroblasts (data not shown). MMP7 was expressed rarely in some macrophages (marked by CD163) and epithelial cells **(Supplementary Table 6, Figure 4D, Supplementary Figure 13)**.

We also stained several other proteins that were identified by SOMAscan. IL-18 staining was limited in non-ILD controls but increased within macrophages, inflammatory cells, and epithelial cells in both sJIA and other ILD cases. In most biopsies, galectin-3 was expressed strongly in alveolar macrophages and more mildly in bronchial and alveolar epithelial cells. Robust expression of CCL2 was observed in the presence of neutrophilic and monocytic inflammation. Rare monocytes and macrophages expressed CCL17 in sJIA cases. **(Supplementary Table 6 and data not shown)**

### Validation of ICAM5 as a biomarker for lung diseases in sJIA

We next organized an independent multi-center validation cohort consisting of 49 serum samples and 29 plasma samples from healthy controls, inactive sJIA, active sJIA, MAS, sJIA-PAP **(Supplementary Table 7)**. Reviewing the validation cohort for intercurrent lung disease identified a few sJIA patients with transient, non-PAP lung ailments (e.g., lobar pneumonia). These we classified as sJIA patients with “other lung disease” and included due to the association of ICAM5 with lung disease in contexts beyond sJIA-PAP **(Supplementary Table 1)**. The validation cohort samples were assayed for IL-18, CXCL9, ICAM5, and MMP7 by antibody-based methods. Serum and plasma samples showed similar trends and dynamic ranges. Compared with healthy controls and inactive sJIA, the levels of IL-18 were elevated in active sJIA, MAS and sJIA-PAP **(Figure 5A)**, and levels of CXCL9 were elevated in many MAS patients **(Figure 5B)**. In contrast, MMP7 and ICAM5 were more specifically and significantly increased in the sJIA-PAP group **(Figure 5C-D)**. Consistent with its expression in lung, these findings support ICAM5 as a potential biomarker for lung disease in sJIA **(Figure 5E)**, distinct from the inflammatory changes associated with sJIA/MAS.

**Figure 5:**
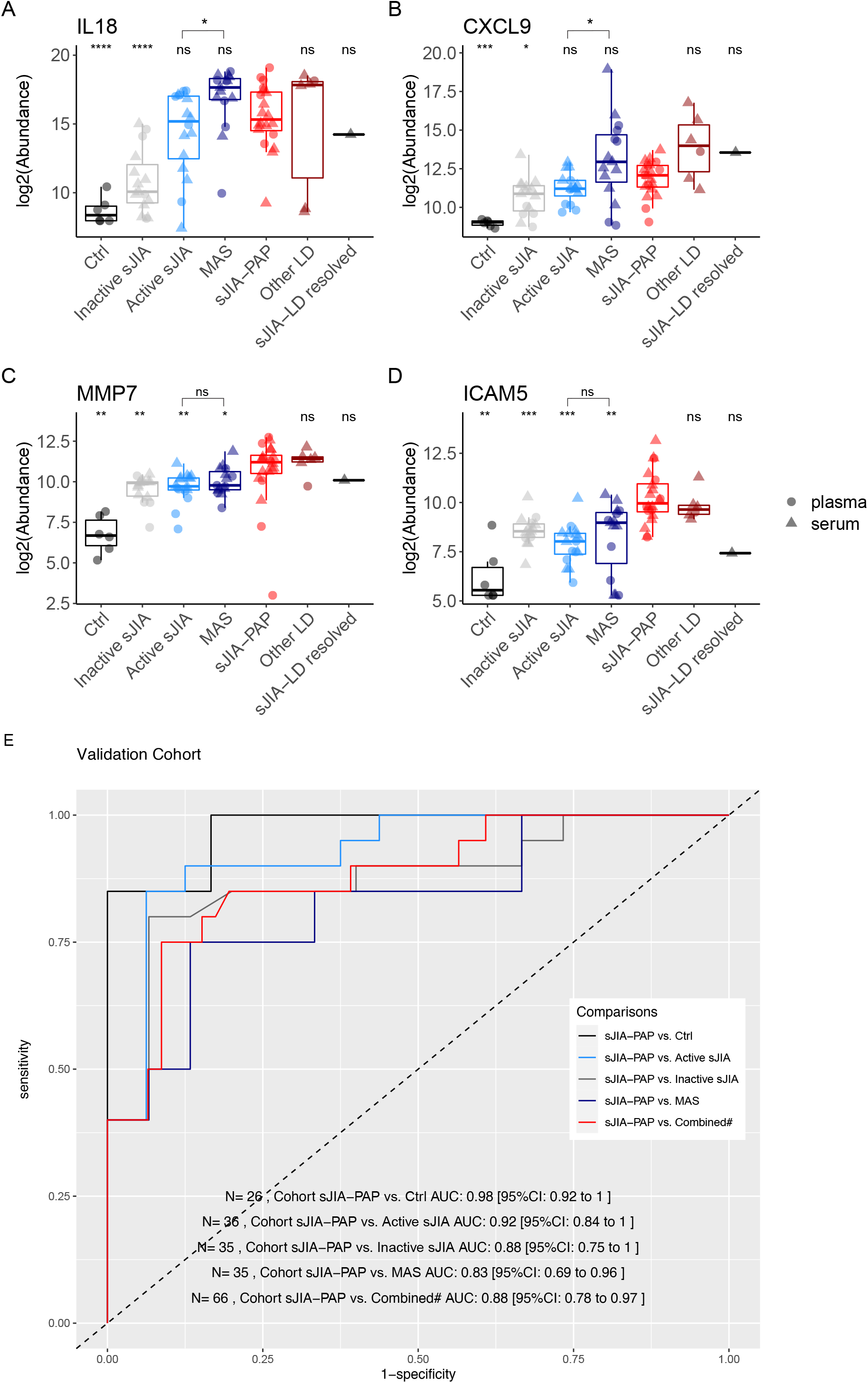
Validation of ICAM-5 as an MAS-independent marker of lung disease in sJIA. Serum (circles) and plasma (triangles) samples from an independent cohort were assayed for IL-18 (A) and CXCL9 (B) by Luminex and for MMP7 (C) and ICAM5 (D) by ELISA. “Other LD” indicates sJIA patients with intercurrent non-PAP lung disease (lobar pneumonia or pulmonary hypertension), whereas “sJIA-LD resolved” indicates a patient with a distant history of radiographic abnormalities resolved at time of sampling. Further details in **Supplemental Tables 1 & 7**. Between-group comparisons were performed using Wilcoxon signed-rank test without the assumption of normal distribution, and reflect comparisons to the sJIA-PAP group unless otherwise indicated. *, p<0.05; **, p<0.01; ***, p<0.001; ****, p< 0.0001. (E) Receiver Operating Characteristic (ROC) analyses of ICAM-5 distinguishing sJIA-PAP from other forms of sJIA. # the combined included inactive/active sJIA and MAS.

## Discussion

Serum proteome profiles can provide clues to pathogenesis as well as clinically actionable biomarkers and can be particularly useful to investigate novel complications of diseases, such as PAP/ELP arising in sJIA patients. To approach both biological and clinical questions concerning sJIA and its serious complications, MAS and PAP, we measured serum levels of 1271 proteins using SOMAscan, verified selected data from this platform using an antibody-based approach and, importantly, validated key results in an independent cohort. Our analyses yielded several insights. First, they provided an unbiased confirmation of known biomarkers of sJIA (S100 proteins, SAA, CRP, IL-6) and MAS (ferritin, IL-18) and also corroborated the more recent finding of elevated IL-18 in sJIA-PAP cases[8, 9, 18, 19]. Second, they identified new proteins/pathways of potential utility in understanding sJIA and MAS, including glycolytic enzymes and heat shock proteins that can act as extracellular DAMPs. Correlation analyses suggested that the sJIA and MAS serum proteomes were related. Finally, the proteins associated with sJIA-PAP reflected inflammation programs distinct from sJIA/MAS. Immunostaining identified cells in lung that may be important sources of serum proteins associated with sJIA-PAP. We validated ICAM5 as a biomarker of lung disease in an independent set of sJIA samples, most usefully in association with sJIA-PAP.

Among the novel proteins identified in the MAS profile, the HSPs may reflect an unfolded protein response, such as that reported to sustain macrophage survival in atherosclerotic lesions[20]. However, these chaperone proteins also play extracellular roles in wound healing, tissue regeneration, and immune responses[21]. Their elevated serum concentration may reflect stress-induced secretion or cell death[22]. Inflammatory forms of cell death (pyroptosis, necroptosis, etc.) are capable of releasing alarmins and DAMPs, such as HSPs, S100 proteins, and IL-33.

In addition, glycolysis-associated proteins **(Figures 2B, D)** were associated with the sJIA and MAS components. Increasingly, aerobic glycolysis is recognized as a necessary metabolic state to execute inflammatory programs in many immune cell types. Animal studies suggest inhibiting glycolysis may be therapeutic in cytokine storms[23]. Many glycolytic enzymes “moonlight” as regulators of inflammatory responses[24-26]. The sJIA and MAS serum programs also included many neutrophil/monocyte proteins (e.g., S100 proteins, PR3, MPO, lipocalin-2, CD163, CD177, **Supplementary Tables 2 and 3**), possibly reflecting their secretion and/or release during cell death.

A high frequency of MAS in sJIA-PAP[7, 8] suggested a connection between sJIA-PAP and the IL-18-interferon gamma (IFNγ) axis underlying MAS. Indeed, sJIA-PAP patients’ peripheral blood often carries an IFN transcriptional signature[9]. In our data, a portion of sJIA-PAP sera (sJIA-PAP^FCHi^) showed elevated CXCL9 and CXCL10 at levels similar to those of the MAS group, although CXCL9 and CXCL10 were not consistently elevated in a previous analysis of bronchoalveolar lavage (BAL) fluid from six sJIA-PAP patients[8]. Combined, these data could be consistent with a role for MAS up- or downstream of PAP **(Supplementary Figure 14)**. Notably, sJIA-PAP was reported in some patients without preceding MAS[7]. We found the sJIA-PAP signature did not correlate with MAS serum activity across patients **(Figures 1D,F)** or timepoints **(Supplementary Figure 7)**, and many cytokines/chemokines had similar abundance in both sJIA-PAP^FCLo^ and sJIA-PAP^FCHi^ groups **(Figure 3B)**. Our data demonstrate that the sJIA-PAP proteome can be present without high MAS activity, supporting the independent origin model **(Supplementary Figure 14B)**. The implication of this model is that treatments targeting the parallel sJIA and MAS proteome patterns (anti-IL-1, anti-IL-6, anti-IFNγ) may be insufficient to manage sJIA-PAP. However, it is also possible that MAS activity is necessary for PAP (either initiation and/or progression), and the sJIA-PAP component indicates lung-specific damage/healing responses rather than a primary pathologic process. Understanding the interaction between the IL-18/IFNγ axis and PAP development will be crucial as therapies targeting this axis become available[27].

A more practical clinical concern is the need for diagnostic and monitoring biomarkers for sJIA-PAP. LDH, surfactant proteins, and MUC-1 (the source of the KL-6 antigen) are biomarkers commonly associated with lung disease [28-32]. Of these, we only observed elevation of LDH in sJIA-PAP **(Supplementary Figure 10)**, and it was also elevated in sJIA and MAS. Our findings in two independent cohorts suggest ICAM5 and MMP7 may serve better to identify lung disease distinct from sJIA/MAS activity.

However, it is unlikely that they are specific to sJIA-PAP. We found elevated MMP7 in autoimmune/hereditary PAP, and elevation of both ICAM5 and MMP7 in a few patients with SAVI-related ILD **(Supplementary Figure 10)**. In our validation cohort, using more available antibody-based methods, ICAM5 consistently and specifically increased in sJIA-PAP, compared to sJIA and MAS. In line with our hypothesis, ICAM5 appeared also to be elevated in cases with pneumonia or pulmonary hypertension, but normal in an sJIA patient with a distant history of lung disease that had resolved at sampling **(Figure 5D, Supplementary Table 1)**. Elevation of ICAM5 was reported in idiopathic pulmonary fibrosis (IPF)[33, 34], rheumatoid arthritis-associated ILD (RA-ILD)[35], and bronchoalveolar lavage (BAL) fluid from neuroendocrine hyperplasia of infancy (NEHI) [36] **(Supplementary Table 8)**.

In previous reports, the tissue or cell of origin for these biomarkers was not studied. We found ICAM5 predominantly in interstitial fibroblasts by immunostaining **(Figure 4D)**, and in fibroblasts, type II alveolar (AT2) cells, and ciliated cells by RNA-sequencing[37] **(Supplementary Figure 15)**. MMP7 has been shown to efficiently cleave ICAM5 and is also elevated in various other ILDs [33, 35, 38]. We hypothesize that ICAM5 elevation in blood may be traced back to lung-specific activity of proteases like MMP7 in sJIA-PAP, and the marker pair may be elevated in a variety of lung diseases.

Cytokines/chemokines contributing to the sJIA-PAP component, particularly CCL17/TARC, CCL7/MCP3, CCL25, GDF15/MIC-1[39] and CCL11/eotaxin-1 **(Supplementary Table 8)**, could be part of pro-fibrotic and/or type 2 immune responses. They, along with MMP7, have been elevated in the proteomic profiles of other ILDs[33-36]. In a mouse model, expression of a PAP-causing surfactant mutant led to overexpression of CCL17, CCL7, and MMP7 proteins in AT2 cells[40]. CCL11 has a profibrotic effect in the lung[41-43]. These sJIA-PAP-associated chemokines also are induced during Type 2 immune responses[44]. Lung biopsies in sJIA-PAP, even from children with long-standing disease, showed remarkably little fibrosis[6-9], possibly due to the young age of the subjects. Previously, we reported atypical rashes (56%) and eosinophilia (37%) during treatment with IL-1 or IL-6 inhibitors in sJIA-PAP[7]. These clinical findings, a minimally-fibrotic lung pathology, recent evidence for a remarkable HLA association[45], and the above serum chemokine elevations (particularly CCL11 and CCL17), are together consistent with a delayed-type drug hypersensitivity reaction[46]. In the current study, we were unable to analyze the impact of IL-1/IL-6 inhibitors on the serum proteome profile, as all the sJIA-PAP patients were under IL-1 or 6 inhibition at sample collection.

This exploratory study has several limitations. First, though the largest to date, our limited sJIA-PAP sample size may not reflect the between-patient heterogeneity or capture the temporal spectrum of this syndrome. Secondly, because all sJIA/MAS/sJIA-PAP patients were being treated at sampling (sometimes with two or more agents), we cannot exclude confounding by disease activity and treatment. However, we previously observed little treatment variation between sJIA-PAP and sJIA patients of comparable disease activity without PAP[7] suggesting small between-group differences in treatment. Nonetheless, clinical/treatment heterogeneity could contribute to biomarker heterogeneity, such as the variability we observed in levels of CXCL9/10, two chemokines rapidly responsive to treatment[47], in the MAS group **(Figure 3D, Figure 5B)**. More recent population biomarker studies highlight the practical value of validated findings in clinically heterogeneous cohorts, leading to successful translation into point-of-care diagnostics[48, 49].

We have leveraged a novel, high-dimensional proteomics platform to identify serum proteins relevant to sJIA, MAS, and the life-threatening development of lung immunopathology; and we have used complementary techniques to localize and validate the results. Unbiased analyses reinforced the primacy of known biomarkers for sJIA and MAS and highlighted novel markers and pathways. Analysis of sJIA-PAP revealed features of smoldering MAS in many, but also a distinct serum proteome with features of lung-specific inflammation, damage repair, and/or hypersensitivity responses. Further, we identified a biomarker (ICAM5) that may be useful to screen for or monitor lung disease in children for whom functional or radiologic testing may be impractical or high-risk. Prospective, longitudinal studies of biomarkers like ICAM5 in patients with sJIA are warranted to directly test their diagnostic and/or prognostic value. Finally, our data can serve as a resource to investigators, clinicians, and families grappling with the management of sJIA, MAS, and PAP.

## Data Availability

The SOMAscan data will be uploaded to GEO (Gene Expression Omnibus).

## Acknowledgments

The authors are grateful for the assistance of the following: Angelique Biancotto, Katie Stagliano, and Jessica Mann at the NIH Center for Human Immunology. We also thank Bhupinder Nahal and the Division of Pediatric Rheumatology at University of California San Francisco, led by Dr. Emily von Scheven, as well as Dr. Sergio Vargas of the Program in Microbiology, Instituto de Ciencias Biomédicas, Universidad de Chile, for collection of several serum samples and associated clinical data.

## Notes

**Funding:** The SOMAscan assay was supported by a grant to the Intramural Research Program of the NIAID from the Systemic JIA Foundation. AAdJ and RGM were supported by the NIAID intramural research program. GC is an Eli Lilly Fellow of the Life Science Research Foundation. SJ is supported by Dean’s Postdoctoral Fellowship, School of Medicine, Stanford. CM and EDM were supported by NIAMS R01 AR066551, AR061297 and Arthritis Foundation Great West Region Arthritis Center of Excellence; VS and EDM were supported by the Lucille Packard Foundation for Children’s Health and a Childhood Arthritis and Rheumatology Research Alliance/Arthritis Foundation grant. CS and SWC were supported by the RK Mellon Institute for Pediatric Research, NIAID K22 AI123366, and NICHD R01 HD098428. PK is funded in part by the Bill and Melinda Gates Foundation (OPP1113682); NIAID 1U19AI109662, U19AI057229, and 5R01AI125197 grants; Department of Defense contracts W81XWH-18-1-0253 and W81XWH1910235; and the Ralph & Marian Falk Medical Research Trust.

**Competing Interest Statement:** The authors declare no conflicts of interest relevant to the submitted work. VS reports personal fees and grants from Novartis. SC reports research support from AB2Bio, Simcha Therapeutics, and IMMvention Therapeutix. RG-M reports grants from Lilly and SOBI. EM reports grants from Novartis and GlaxoSmithKline. PK reports personal fees from Inflammatix, Inc. and Cepheid. GS reports personal fees from Novartis and SOBI. GD reports personal fees from Novartis. AG reports grants and personal fees from SOBI, Novartis, and AB2Bio.

### Competing Interest Statement

VS reports personal fees from Novartis, grants from Novartis, outside the submitted work. SC reports grants from Systemic JIA Foundation, grants from NICHD, grants from RK Mellon Institute, during the conduct of the study; grants from AB2Bio, Ltd., other from Simcha Therapeutics, grants from IMMvention Therapeutix, outside the submitted work. RG-M reports grants from Systemic JIA Foundation during the conduct of the study; grants from Lilly, grants from SOBI outside the submitted work. EM reports grants from Lucille Packard Foundation for Children Health, grants from CARRA-Arthritis Foundation, during the conduct of the study; grants from Novartis, grants from GSK, outside the submitted work. PK reports personal fees and other from Inflammatix, Inc., personal fees from Cepheid, outside the submitted work. GS reports personal fees from Novartis, personal fees from SOBI, outside the submitted work. GD reports personal fees from Novartis Pharmaceuticals, outside the submitted work. AG reports grants and personal fees from Sobi, grants and personal fees from Novartis, grants and personal fees from AB2Bio, during the conduct of the study.

### Funding Statement

The SOMAscan assay was supported by a grant to the Intramural Research Program of the NIAID from the Systemic JIA Foundation. AAdJ and RGM were supported by the NIAID intramural research program. GC is an Eli Lilly Fellow of the Life Science Research Foundation. SJ is supported by Dean Postdoctoral Fellowship, School of Medicine, Stanford. CM and EDM were supported by NIAMS R01 AR066551, AR061297 and Arthritis Foundation Great West Region Arthritis Center of Excellence; VS and EDM were supported by the Lucille Packard Foundation for Children Health and a Childhood Arthritis and Rheumatology Research Alliance/Arthritis Foundation grant. CS and SWC were supported by the RK Mellon Institute for Pediatric Research, NIAID K22 AI123366, and NICHD R01 HD098428. PK is funded in part by the Bill and Melinda Gates Foundation (OPP1113682); NIAID 1U19AI109662, U19AI057229, and 5R01AI125197 grants; Department of Defense contracts W81XWH-18-1-0253 and W81XWH1910235; and the Ralph & Marian Falk Medical Research Trust.

### Author Declarations

Ethics approval for collection of clinical data and patient serum through institutional review boards at the following institutions: Stanford University, University of Cincinnati, University of Pittsburgh and National Institute of Allergy and Infectious Diseases at the National Institutes of Health.

