## Supplemental figures for "Serum proteome analysis of systemic JIA and related pulmonary alveolar proteinosis identifies distinct inflammatory programs and biomarkers"

### Supplementary Figure 1

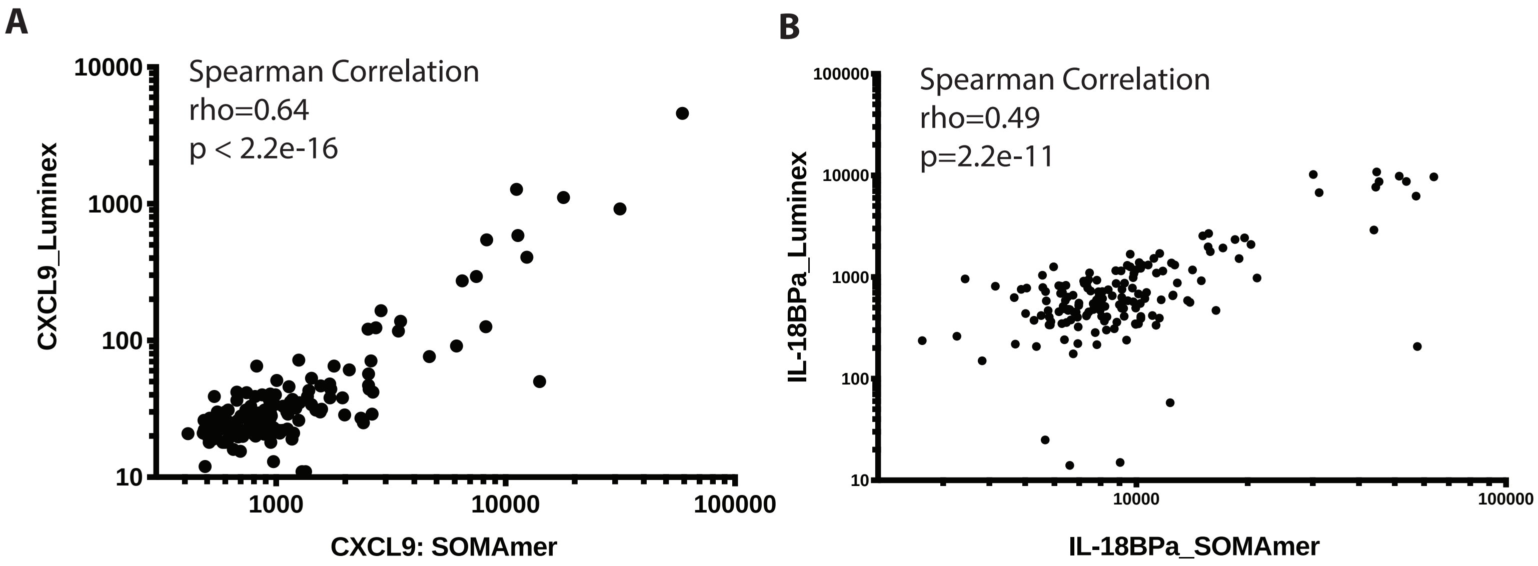


Supplementary Figure 1. Correlation between SOMAscan results and antibody based Luminex results.

### Supplementary Figure 2

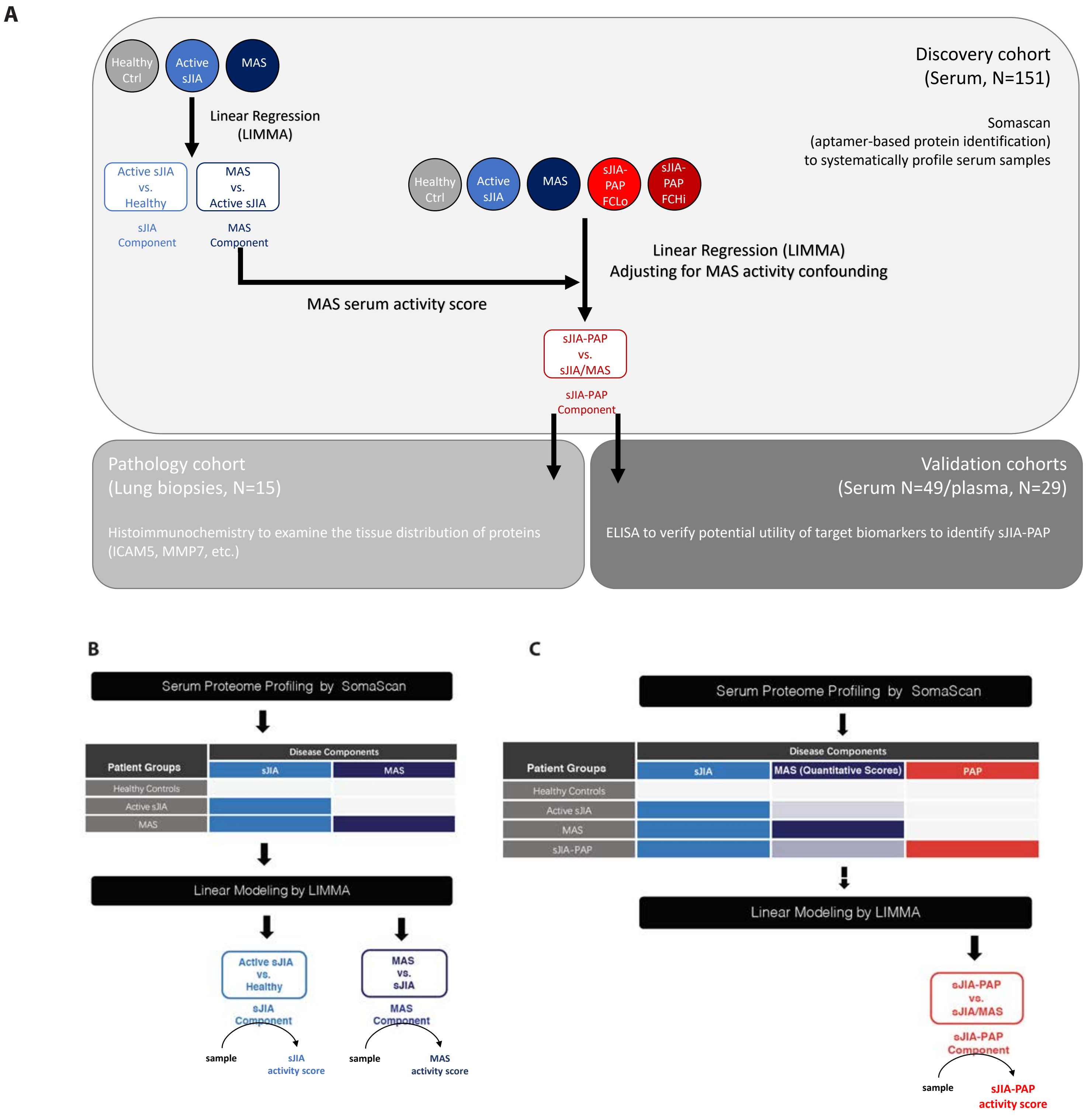


**Analytical scheme of the study.** A, The flowchart of the study. The two limma models are presented in B and C.

### Supplementary Figure 3

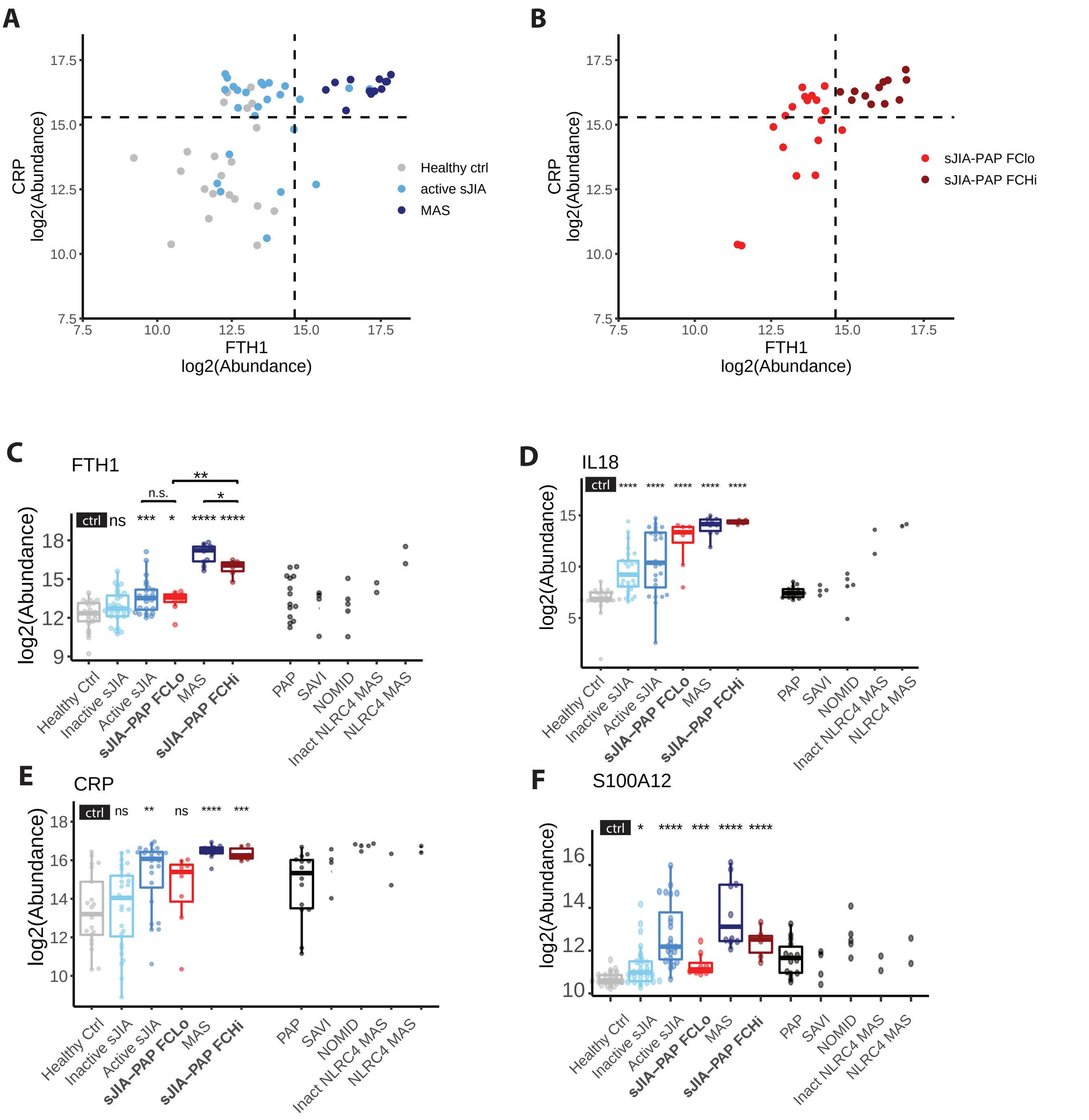


**Known sJIA and MAS biomarker abundance measured by SOMAscan in different disease groups.** A-B, Log2 abundance of ferritin (FTH) and CRP as measured by SOMAscan were used to identify groups with different MAS activity levels. The thresholds used in CRP and ferritin levels to define FCHi and FCLo in the sJIA-PAP patient group are shown in the dashed lines. C-F, The stars denote the significance in the comparison between the designated group and the healthy controls. sJIA/MAS diseases are colored in shades of blue, groups with sJIA-PAP component are colored in shades of red. The between group comparisons were performed using Wilcoxon signed-rank test with p value adjusted by Benjamini-Hochberg Procedure. Unless marked by a bracket, the comparison is between each indicated group and the healthy control group. Comparator groups (black) were not included in statistical analysis due to low sample numbers. *, $p<0.05$; **, $p<0.01$; ***, $p<0.001$; ****, $p< 0.0001$. To facilitate analysis, gene names are presented but represent protein targets, see online supplemental methods.

### Supplementary Figure 4

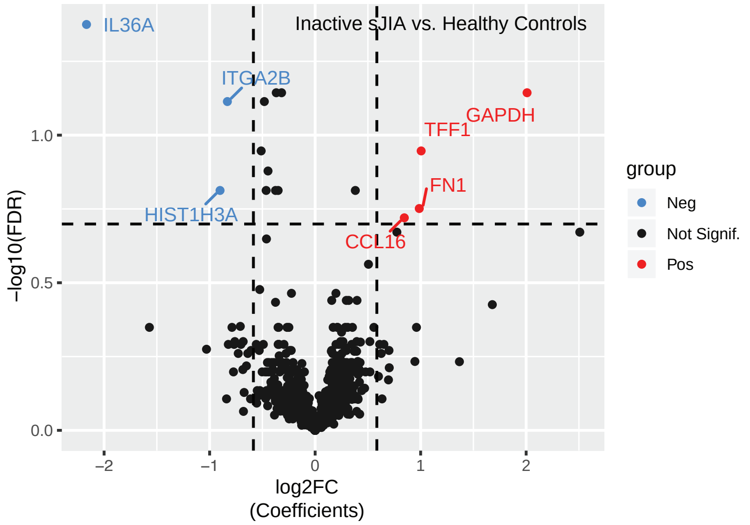


**The volcano plot for the inactive sJIA component (inactive sJIA vs. healthy controls).** Significance thresholds are represented by the dashed lines: False Discovery Rate (FDR, adjusted p value) < 0.2 and absolute fold change > 1.5. To facilitate analysis, gene names are presented but represent protein targets, see online supplemental methods.

### Supplementary Figure 5

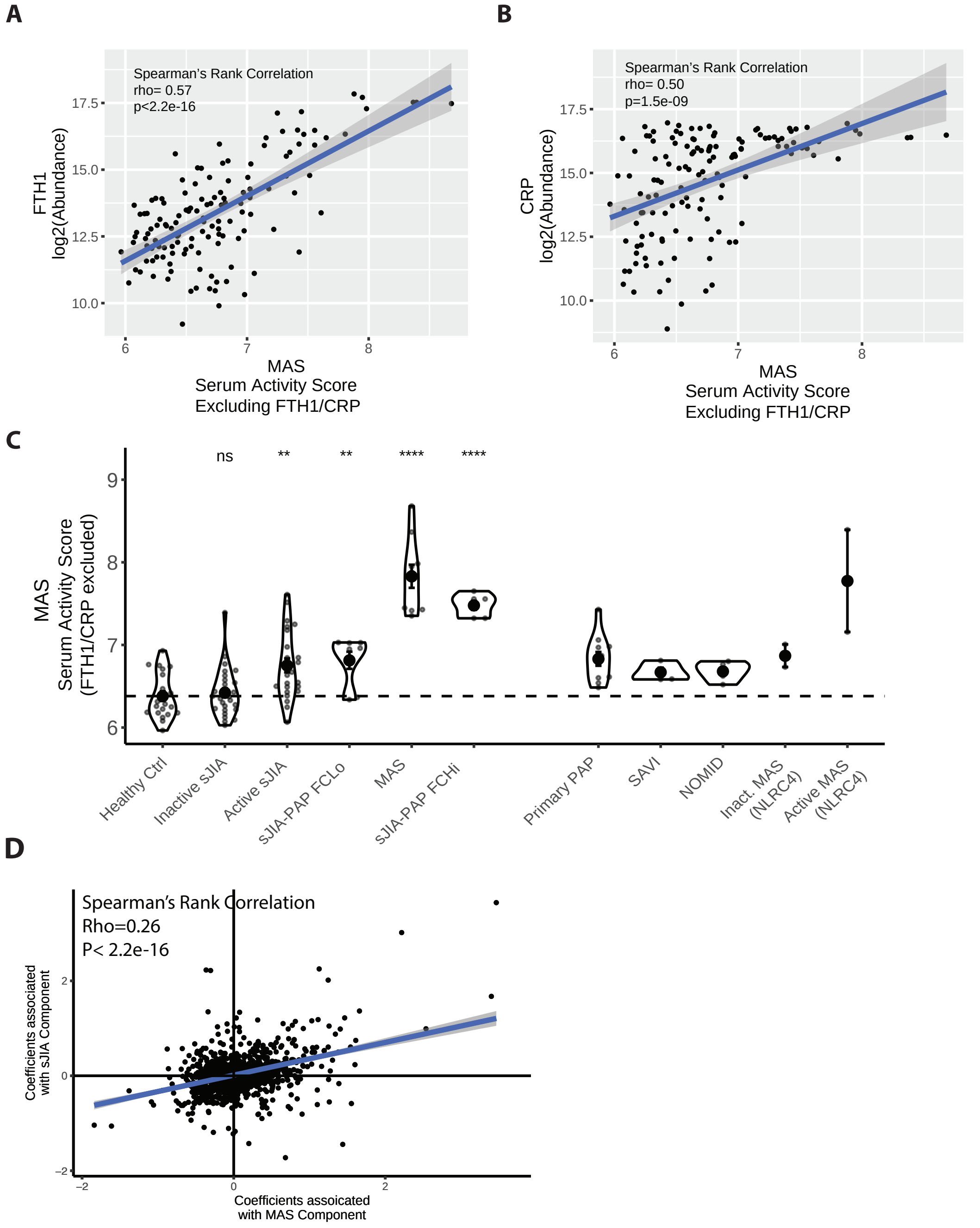


**Measuring MAS activity in sJIA-PAP.** To capture the overall abundance change, we calculated a MAS serum activity score using the geometric means of significantly changed proteins (see online supplemental methods). A-B) After removing FTH and CRP from the calculation of MAS serum activity score, the score still correlates with FTH and CRP. C) The MAS serum activity scores for different groups are shown in the boxplot. The between group comparisons, between each indicated group and the healthy control group, were performed using Wilcoxon signed-rank test with p value adjusted by Benjamini-Hochberg Procedure. Comparator groups (on right side of box plots) were not included in statistical analysis due to low sample numbers. D) The correlation between the coefficients, assigned by LIMMA model to the sJIA and MAS disease components, was analyzed by Spearman correlation. P value is shown.

### Supplementary Figure 6

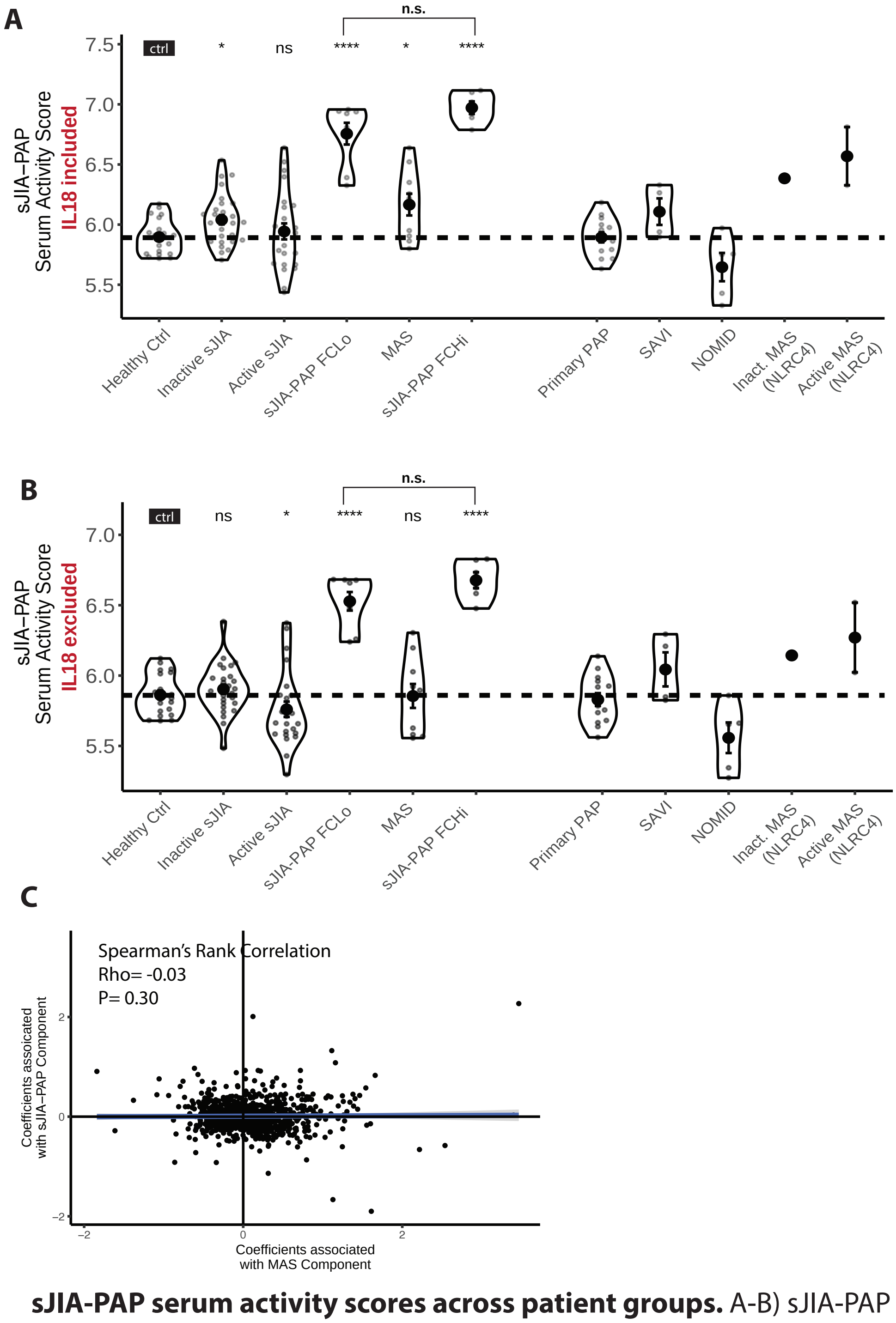


**sJIA-PAP serum activity scores across patient groups.** A-B) sJIA-PAP serum activity scores across different patient groups with and without the contribution of IL-18 (note sJIA-PAP score of the MAS group). The between group comparisons, between each indicated group and the healthy control group, were performed using Wilcoxon signed-rank test with p value adjusted by Benjamini-Hochberg Procedure. Comparator groups were (on right side of box plots) not included in statistical analysis due to low sample numbers. *, $p<0.05$; **, $p<0.01$; ***, $p<0.001$; ****, $p< 0.0001$. C) Shown is the correlation between the coefficients, assigned by LIMMA model, to the MAS and sJIA-PAP disease components, analyzed by Spearman correlation. P value is shown.

### Supplementary Figure 7

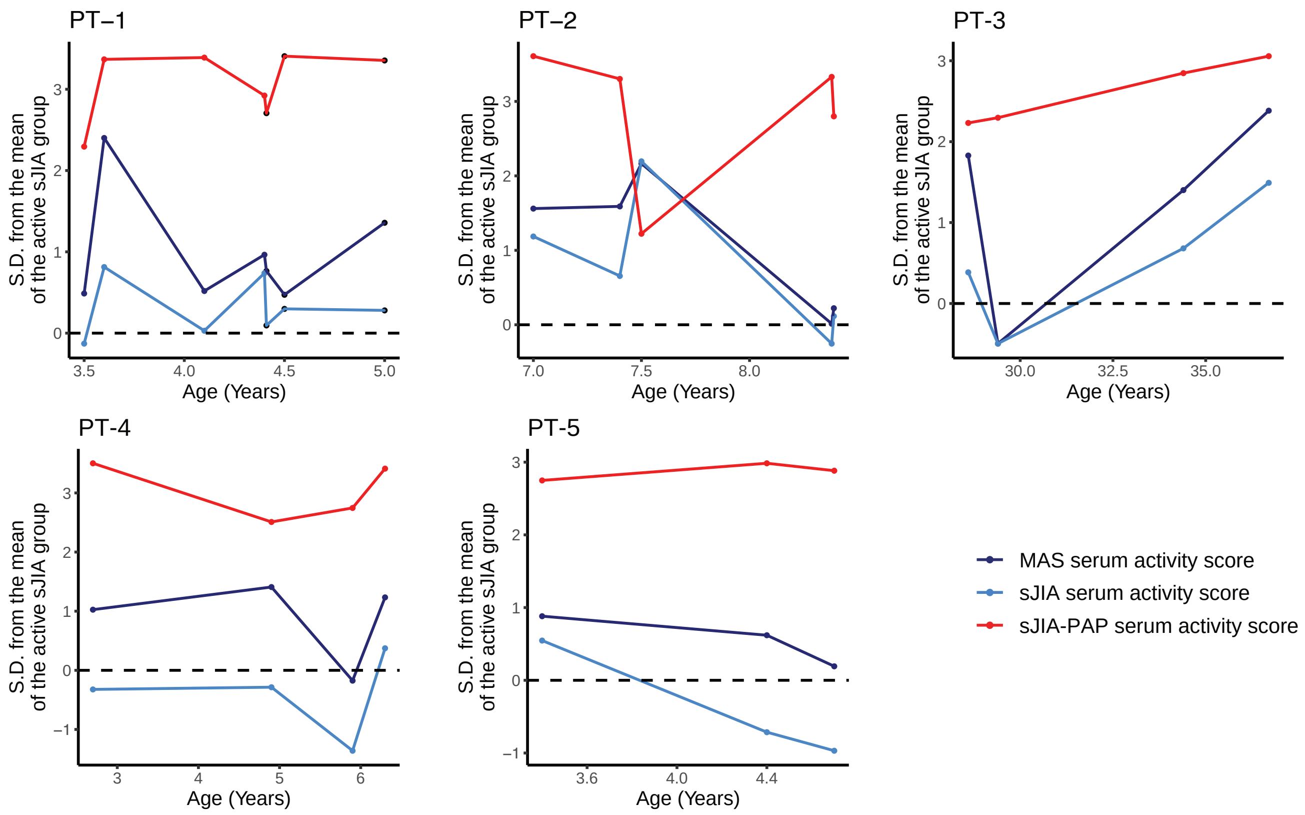


**Longitudinal alterations of disease component-specific serum activity in patients with sJIA-PAP.** The longitudinal profiles of MAS (black), sJIA (blue), and sJIA-PAP (red) serum activity scores are shown. Only patients with 3 or more timepoints are included. Y axis denotes the difference between the measurement in each timepoint and the mean of healthy controls, normalized by the standard deviations in the healthy controls.

### Supplementary Figure 8

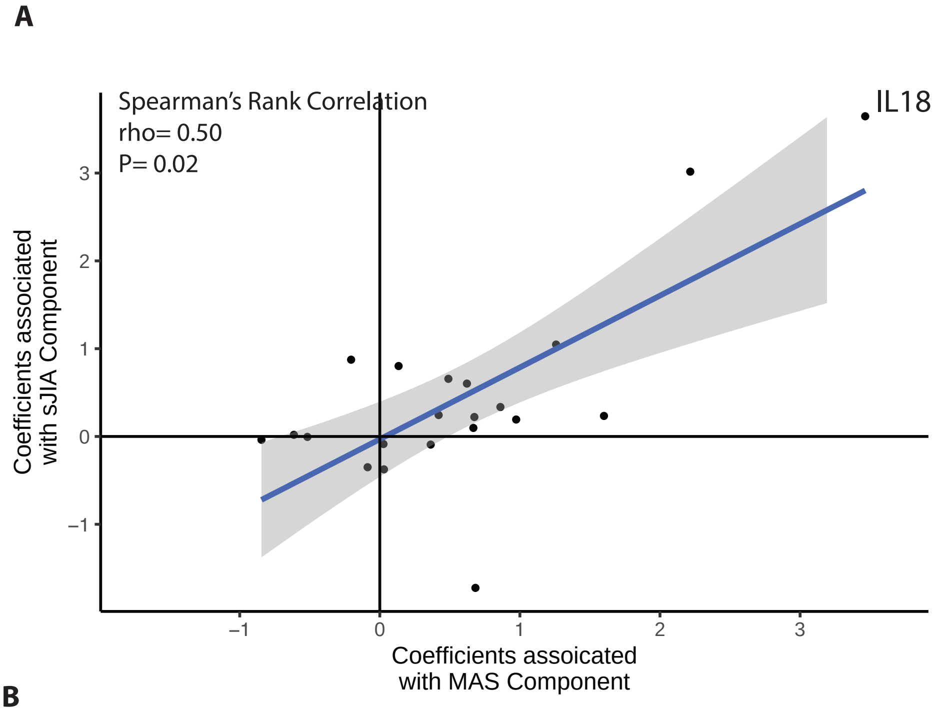


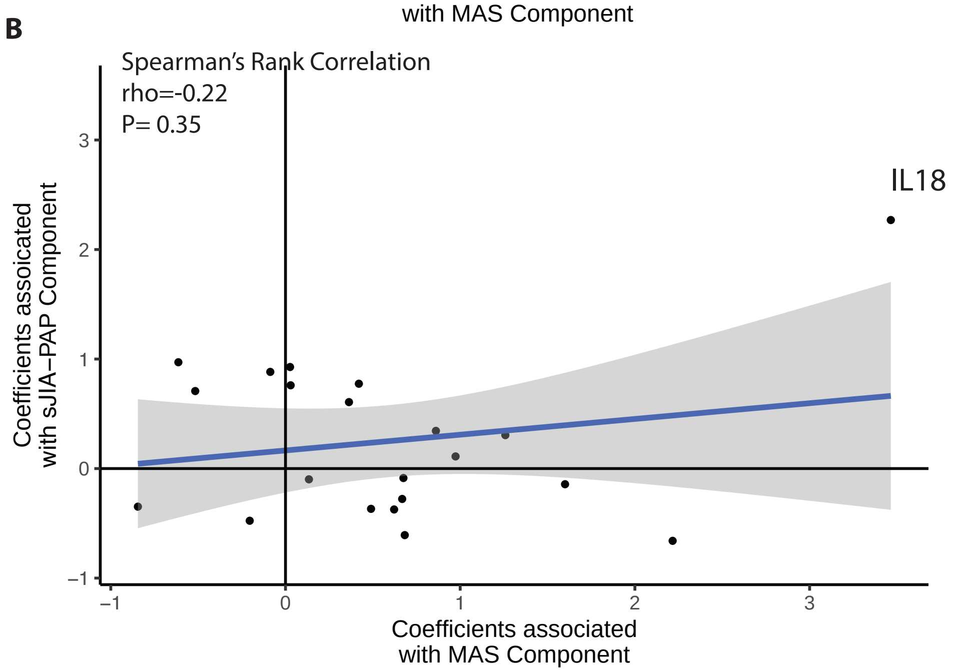


**Comparisons between cytokine/chemokine profiles associated with different disease components.** Included are cytokines and chemokines that were present in the SOMAscan dataset, were included in the GO terms GO:0005125: cytokine activity or GO:0042056: chemoattractant activity and were significantly altered in at least one of the three disease components (sJIA, MAS, sJIA-PAP). The correlation between the coefficients, assigned by LIMMA model, to each disease component is shown. (A, B). Each dot represents a cytokine/chemokine. P value is shown.

### Supplementary Figure 9

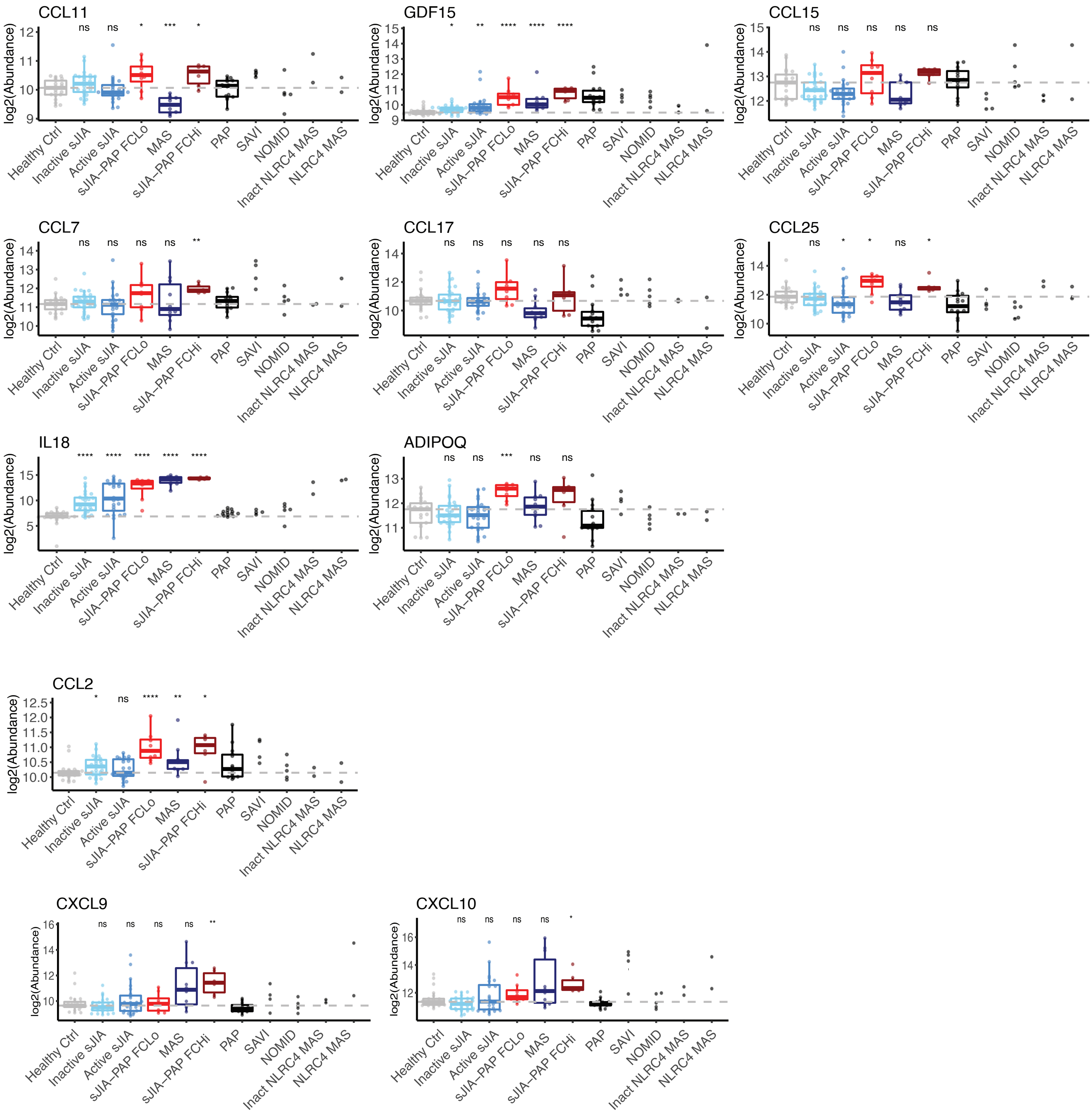


**Levels of cytokines and chemokines significantly associated with the sJIA-PAP disease component.** Note: CCL2 did not reach pre-defined significance threshold (figure 2B), but it is included due to its potential functional relevance. CXCL9 and CXCL10, two interferon inducible chemokines, were not associated with the sJIA-PAP disease components (see online supplemental table S4) but are shown here to examine the relationship between sJIA-PAP and the interferon response. The between group comparisons, between each indicated group and the healthy control group, were performed using Wilcoxon signed-rank test with p value adjusted by Benjamini-Hochberg Procedure. Comparator groups (black) were not included in statistical analysis due to low sample numbers. *, p<0.05; **, p<0.01; ***, p<0.001; ****, p< 0.0001. To facilitate analysis, gene names are presented but represent protein targets, see online supplemental methods.

### Supplementary Figure 10

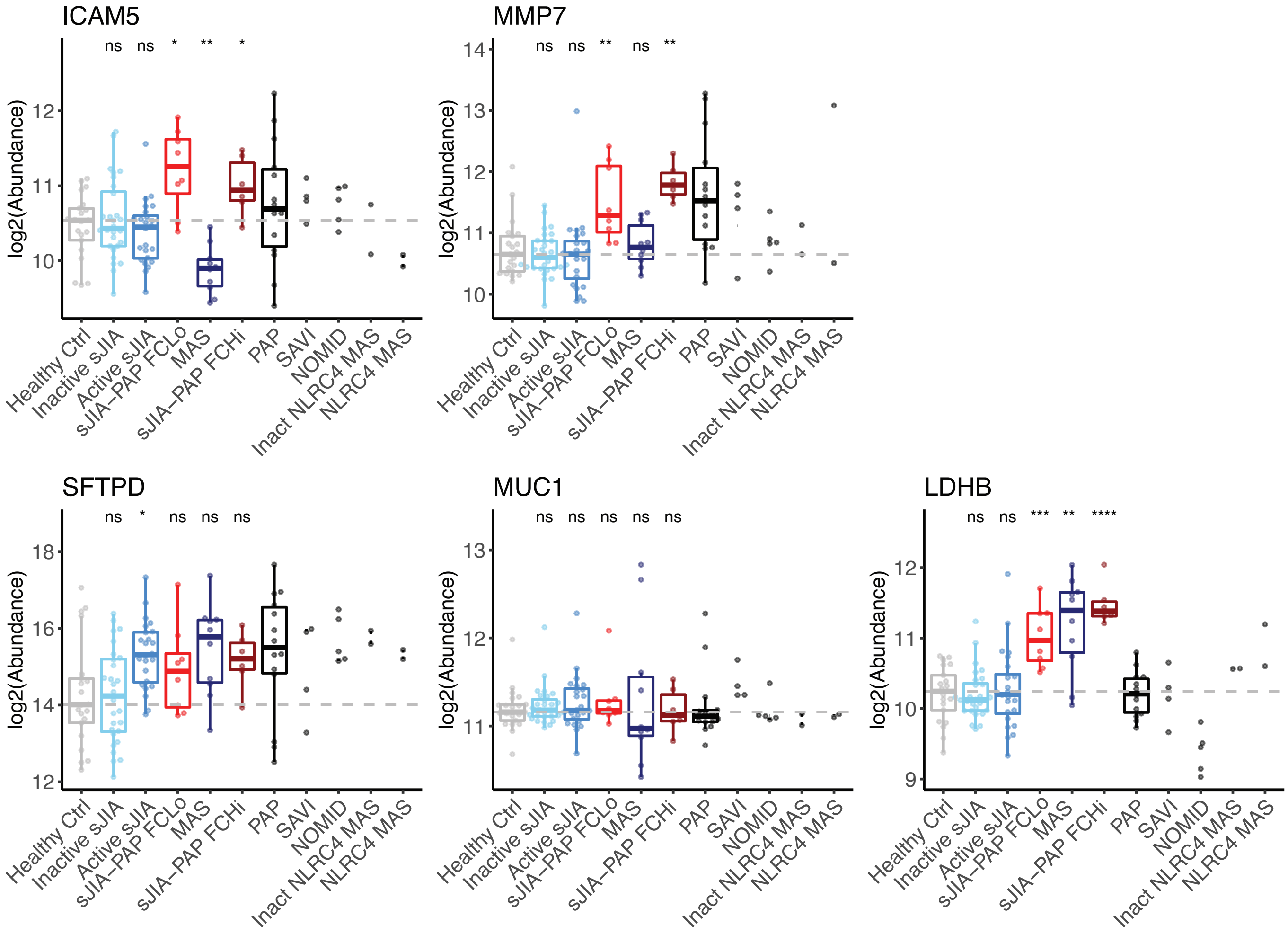


**Levels of biomarkers associated with sJIA-PAP and other lung diseases.** ICAM5 and MMP7 from our analysis are shown alongside other biomarkers related to interstitial lung diseases. SFTPD is surfactant protein D. MUC1 is the protein that contains the KL-6 antigen. The between group comparisons, between each indicated group and the healthy control group, were performed using Wilcoxon signed-rank test with p value adjusted by Benjamini-Hochberg Procedure. Comparator groups (black) were not included in statistical analysis due to low sample numbers. *, $p<0.05$; **, $p<0.01$; ***, $p<0.001$; ****, $p< 0.0001$. To facilitate analysis, gene names are presented but represent protein targets, see online supplemental methods.

### Supplementary Figure 11

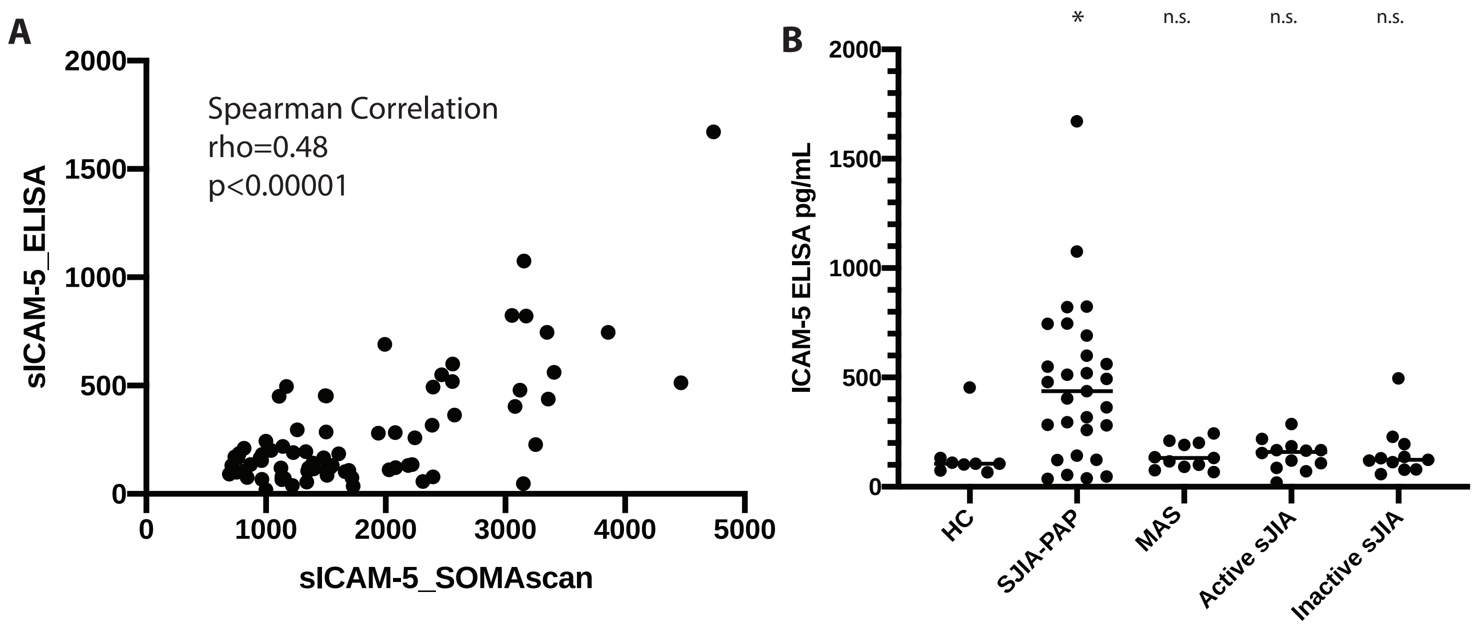


**ICAM5 abundance in the discovery cohort measured by antibody-based ELISA.**
A) The results by ELISA correlated with the SOMAscan. B) The abundance of ICAM5 as measured by ELISA is shown across different patient groups. The between group comparisons, between each indicated group and the healthy control group, were performed using Wilcoxon signed-rank test with p value adjusted by Benjamini-Hochberg Procedure. *, p<0.05; **, p<0.01; ***, p<0.001; ****, p< 0.0001.

### Supplementary Figure 12

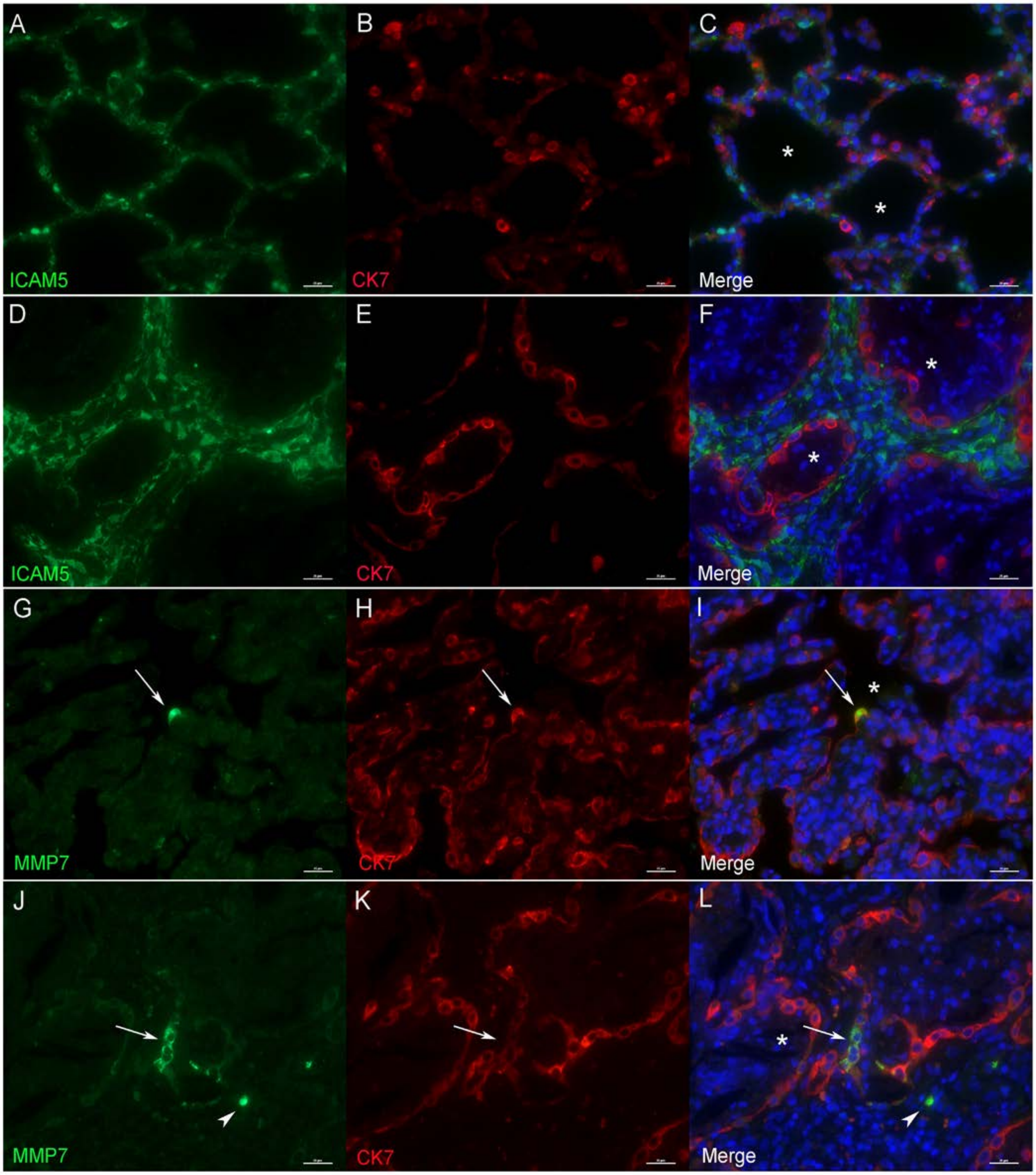


**Tissue disribution of sJIA-PAP biomarkers identified in the serum.** Immunofluorescence for ICAM (A-F) and MMP7 (G-L) with the epithelial marker cytokeratin 7 (CK7) in control lungs (A-C; G-I) and sJIA lungs (D-F; J-L). No colocalization is seen with ICAM5, which is restricted to fibroblasts in the alveolar septa (* denotes alveolar lumen), versus MMP7, which colocalizes with CK7 in rare to few epithelial cells (arrows). Arrowhead shows positive MMP7 in a hematopoietic cell. Nuclei counterstained with DAPI (blue).

### Supplementary Figure 13

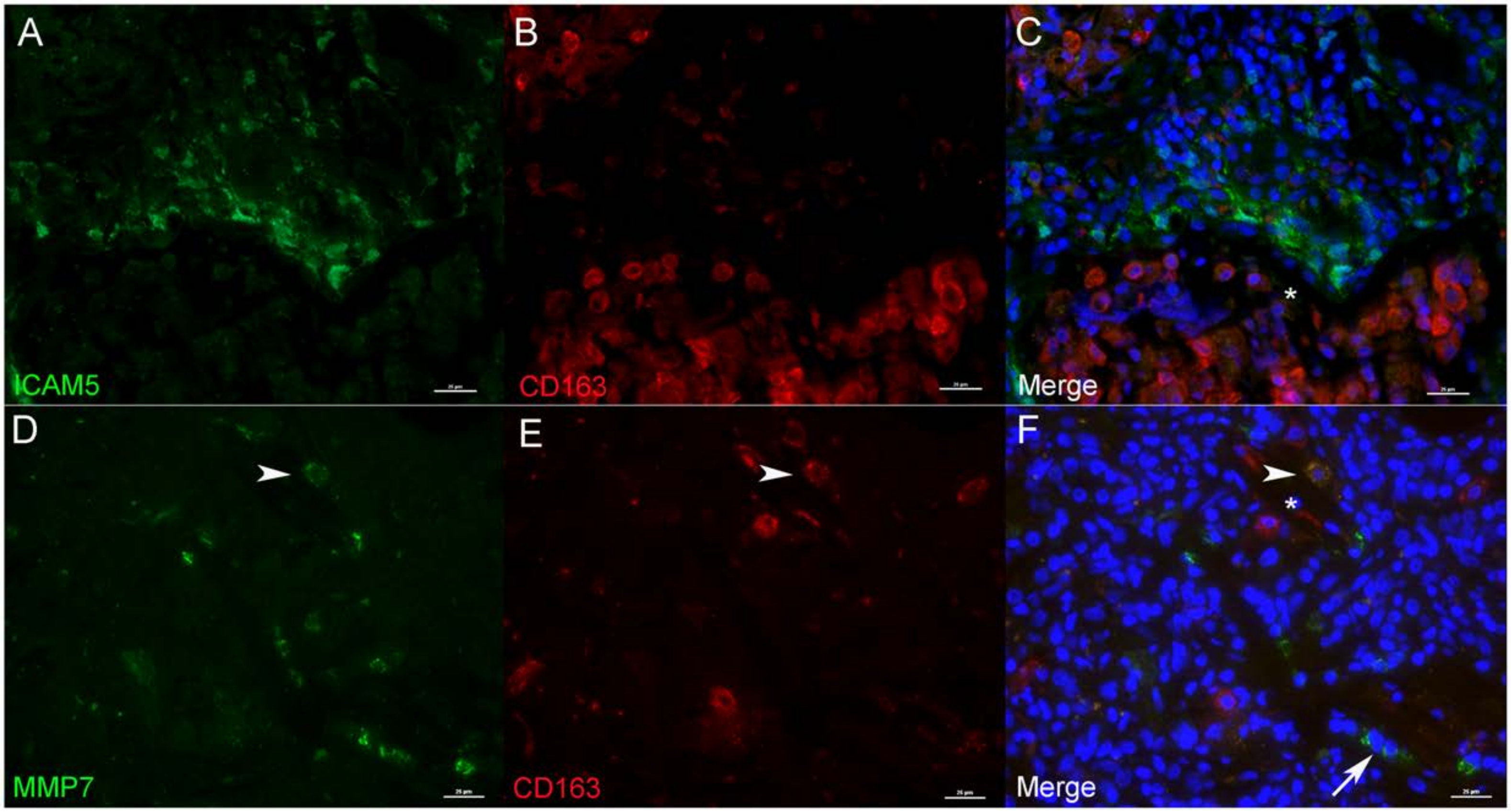


**Immunohistochemistry of lung biopsies from the control and sJIA-PAP cases**
mmunofluorescence for ICAM5 (A-F) and MMP7 (G-L) and the macrophage marker CD163 in control lungs (A-C; G-I) and sJIA lungs (D-F; J-L). No colocalization is seen with ICAM5, which is restricted to expanded fibroblasts in the alveolar septa (* denotes alveolar lumen). MMP7 in rare macrophages colocalized with CD163 (arrowhead); arrow indicates positive MMP7 in epithelial cells. Nuclei counter-stained with DAPI (blue).

### Supplementary Figure 14

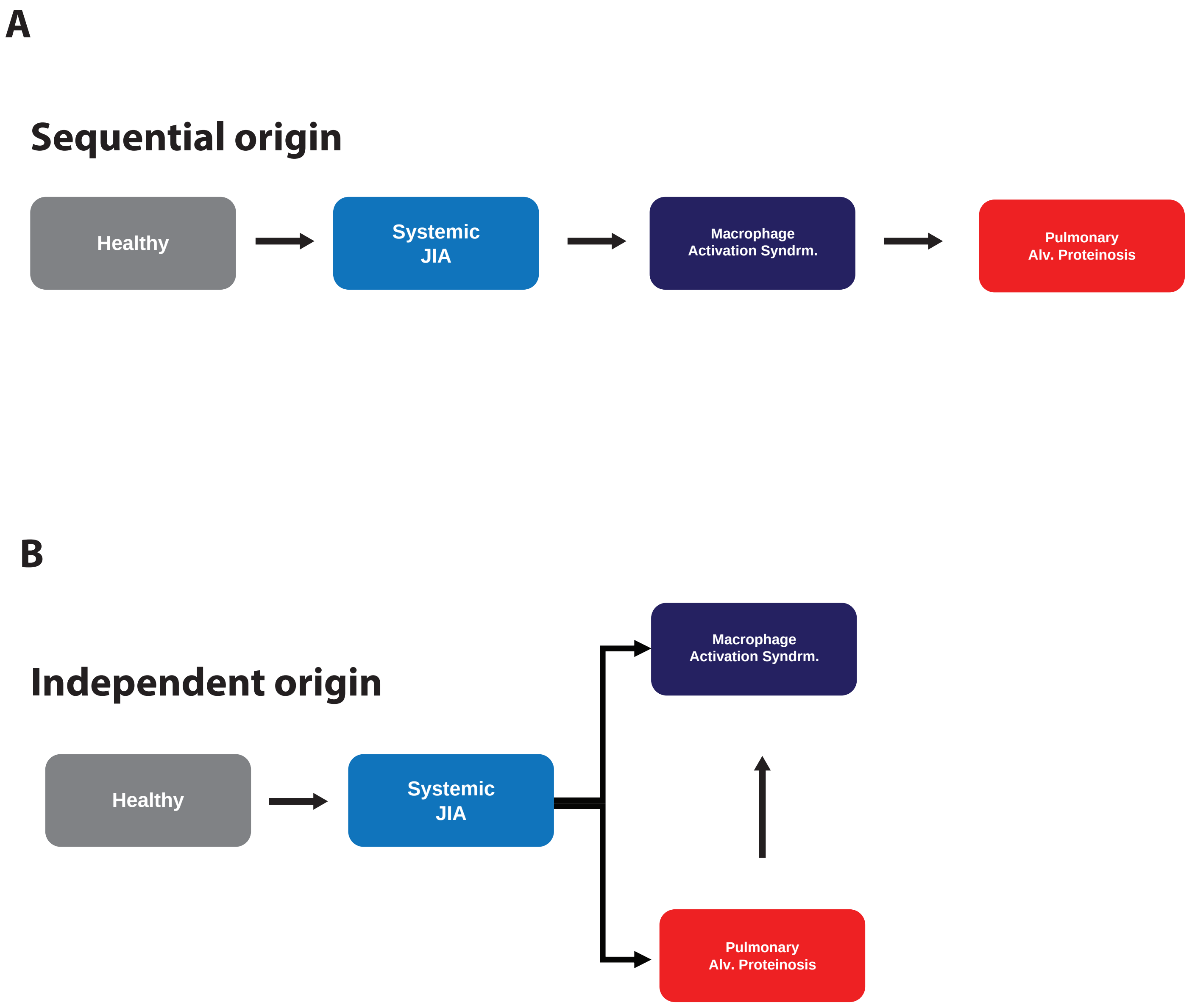


**Two hypothetical models to describe the relationships between sJIA, MAS and sJIA-PAP**
A) The sequential development model: sJIA-PAP results from elevated MAS activities, with shared etiology. B) The independent origin model: sJIA-PAP and MAS are triggered by different underlying causes. In a flare of sJIA-PAP, MAS can be triggered, possibly due to elevated cytokines, such as IL-18.

### Supplementary Figure 15

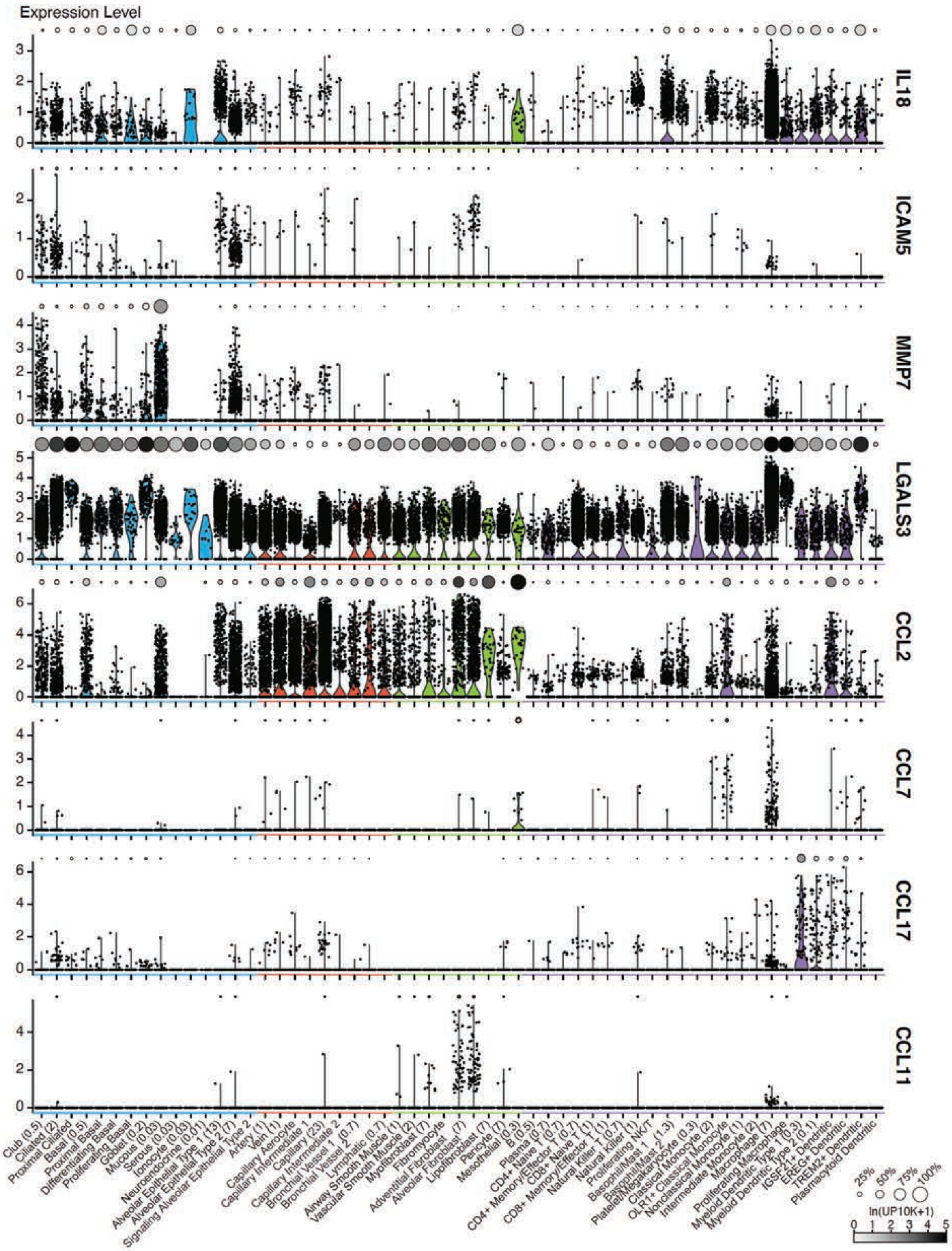


**Expression level (ln(UP10K+1)) of sJIA-PAP signature genes in 57 normal human lung cell types.** Dot plot (color intensity of the dot reflects the mean level of normalized expression and the size reflects the percentage of cells expressing the gene) for each gene is shown above individual violin plots, which show the distribution of the gene expression (each point representing a single cell). Cell types are color coded by compartment (epithelial – blue; endothelial – red; stromal – green; and immune – purple). If reported, relative abundances of the cell types (in percentage) are indicated inside a bracket on the x-axis1. UP10K, UMIs per 10,000.
