## supplemental tables for "Serum proteome analysis of systemic JIA and related pulmonary alveolar proteinosis identifies distinct inflammatory programs and biomarkers"

| Supplementary Table 1: Pulmonary Characteristics of SJIA-PAP patients | | | | | | |
|---|---|---|---|---|---|---|
| Pt ID | clubbing | RadPattern | Bx | Published | Biologic Exposure^ | Current Status |
| **SOMAscan cohort** | | | | | | |
| A | y | ST, GGO, consolidation | PAP/ELP | 1,2 | 1,6 | stable/ongoing |
| B | y | ST, nodules | na | no | 1 | lost to follow-up |
| C | n | GGO -> ST | na | 1,2 | 6 | stable/ongoing |
| D | y | ST, GGO, consolidation | PAP/ELP | 1 | 1 | improved |
| E | y | ST, GGO, consolidation | na | 1 | 1 | improved |
| F* | y | ST, GGO | PAP/ELP | 1 | 1 | deceased |
| G | y | ST, GGO -> consolidation | na -> PAP/ELP | 1 | 1 | stable/ongoing |
| H | y | ST, GGO, consolidation | Likely PAP/ELP | 1 | 1 | stable/ongoing |
| I | y | ST, GGO | na | 1,2 | 1 | stable/ongoing |
| J* | y | ST, GGO, consolidation | PAP/ELP | 1 | 1,6 | deceased |
| **Validation Cohort** | | | | | | |
| A | y | ST, GGO, consolidation | PAP/ELP | 1,2 | 1,6 | stable/ongoing |
| D | y | ST, GGO, consolidation | PAP/ELP | 1 | 1,6 | improved |
| F* | y | ST, GGO | PAP/ELP | 1 | 1 | deceased |
| G | y | ST, GGO -> consolidation | PAP/ELP | 1 | 1 | stable/ongoing |
| K | y | perihilar consolidation # | na | na | 1 | stable/ongoing |
| L | n | ST# | PAP/ELP | 2 | 1,6 | improved |
| M | y | fibrosis, consolidation# | PAP/ELP | 2 | 1,6 | progressive |
| N | y | ST# | PAP/ELP | 2 | 1,6 | stable/ongoing |
| O | y | ST, consolidation# | na | 2 | 1,6 | resolved |
| P | y | ST# | na | na | 1 | stable/ongoing |
| Q | y | ST, consolidation# | PAP/ELP | 2 | 1,6 | stable/ongoing |
| R | y | ST, GGO, consolidation# | PAP/ELP | 2 | 1 | stable/ongoing with tracheostomy |
| S | n | ST, GGO# | na | 2 | 1,6 | stable/ongoing |
| T | y | ST# | na | 2 | 1 | stable/ongoing on oxygen |
| U | y | ST# | PAP/ELP | 2 | 1,6 | progressive |
| V | n | GGO, consolidation# | na | na | 1,6 | stable/ongoing on oxygen |
| E | y | ST, GGO, consolidation# | na | 1 | 1 | improved |
| X | y | ST, nodules, PHTN# | na | na | 1 | stable |
| Y | y | GGO, PHTN# | unknown | na | 1 | stable |
| Z* | y | ST,GGO# | PAP/ELP | na | 1,6 | deceased |
| AA | y | ST,GGO,consolidation# | na | na | 1 | stable/ongoing |
| BB | y | ST# | na | na | 1,6 | stable/ongoing |
| **Other LD** | | | | | | |
| | n | ST, PHTN# | na | na | 1 | no lung disease |
| | n | consolidation# | na | na | 1 | no lung disease |
| | n | consolidation# | na | na | 1 | no lung disease |
| **sJIA-LD resolved** | | | | | | |
| | y | ST, consolidation# | na | 2 | 1,6 | no lung disease |
| ST: multi-lobular intra-lobular septal thickening; GGO=Ground Glass Opacities; PHTN=evidence of pulmonary hypertension such as enlarged pulmonary arteries | | | | | | |
| -> indicates development after sample | | | | | | |
| *Now deceased | | | | | | |
| ^Prior or current exposure to IL-1 blockade (1) or IL-6 blockade (6) | | | | | | |
| #not part of a systematic radiology review | | | | | | |
| (1) Saper, Chen, et al., Ann Rheum Dis, 2019; (2) Schulert et al., Arth Rheum, 2019 | | | | | | |

**Table S2: Proteins whose concentration was significantly altered in the sJIA disease component**

| Gene Symbol ID | Coefficients | t | P.Value | FDR | Full gene name | Prt/Prt Complex mapped* |
|---|---|---|---|---|---|---|
| S100A12 | 2.251266229 | 6.032033241 | 8.58E-08 | 0.000108996 | S100 calcium binding protein A12 | |
| MMP3 | 2.217432084 | 5.546202576 | 5.76E-07 | 0.000243729 | matrix metallopeptidase 3 | |
| PAPPA | 1.175756804 | 5.570455678 | 5.25E-07 | 0.000243729 | pappalysin 1 | |
| S100A9 | 1.099559551 | 5.472243281 | 7.67E-07 | 0.000243729 | S100 calcium binding protein A9 | |
| MCL1 | 0.762550308 | 4.978906915 | 5.00E-06 | 0.001270632 | MCL1, BCL2 family apoptosis regulator | |
| MAPK14 | 0.877587068 | 4.929042672 | 6.02E-06 | 0.001275133 | mitogen-activated protein kinase 14 | |
| CKM | -1.170378828 | -4.853666572 | 7.96E-06 | 0.001445508 | creatine kinase, M-type | |
| CNTN2 | -0.667496825 | -4.710058605 | 1.35E-05 | 0.00214414 | contactin 2 | |
| EDAR | -1.10782229 | -4.612868542 | 1.92E-05 | 0.002442547 | ectodysplasin A receptor | |
| NAGK | 1.364851621 | 4.635406218 | 1.77E-05 | 0.002442547 | N-acetylglucosamine kinase | |
| APOB | -1.426212002 | -4.576739369 | 2.19E-05 | 0.002484366 | apolipoprotein B | |
| HAPLN1 | -1.190719311 | -4.557677813 | 2.35E-05 | 0.002484366 | hyaluronan and proteoglycan link protein 1 | |
| CKB | -1.222006538 | -4.502845551 | 2.86E-05 | 0.002484935 | creatine kinase B | CKB.CKM, CKB |
| FAP | -0.804764545 | -4.480367537 | 3.10E-05 | 0.002484935 | fibroblast activation protein alpha | |
| GPI | 1.047687369 | 4.472551881 | 3.18E-05 | 0.002484935 | glucose-6-phosphate isomerase | |
| IL18 | 3.6475659 | 4.444409295 | 3.52E-05 | 0.002484935 | interleukin 18 | |
| MMP1 | 1.347909655 | 4.459339234 | 3.34E-05 | 0.002484935 | matrix metallopeptidase 1 | |
| GAPDH | 2.228423265 | 4.421293608 | 3.82E-05 | 0.002555909 | glyceraldehyde-3-phosphate dehydrogenase | |
| MAPK3 | 0.78186304 | 4.369730076 | 4.59E-05 | 0.002914891 | mitogen-activated protein kinase 3 | |
| SAA1 | 3.016776649 | 4.303379627 | 5.79E-05 | 0.00350676 | serum amyloid A1 | |
| GPC3 | -0.682791531 | -4.21907112 | 7.78E-05 | 0.003813042 | glypican 3 | |
| LILRB2 | 0.722365392 | 4.18704467 | 8.69E-05 | 0.004063684 | leukocyte immunoglobulin like receptor B2 | |
| FTH1 | 1.673096855 | 4.121412572 | 0.000109005 | 0.004618174 | ferritin heavy chain 1 | FTH1.FTL |
| FUT5 | 0.68456594 | 4.056898669 | 0.000135942 | 0.005380091 | fucosyltransferase 5 | |
| CD109 | -0.586777073 | -3.966380707 | 0.000184769 | 0.006347071 | CD109 molecule | |
| HK2 | 1.218236915 | 3.969635951 | 0.000182752 | 0.006347071 | hexokinase 2 | |
| GSK3A | 0.64139195 | 3.952508008 | 0.000193606 | 0.006475622 | glycogen synthase kinase 3 alpha | GSK3A.GSK3B |
| SLITRK5 | -0.935087283 | -3.919580413 | 0.000216238 | 0.006703381 | SLIT and NTRK like family member 5 | |
| PKM2 | 1.01871023 | 3.883884242 | 0.000243639 | 0.006765746 | NA | |
| PPP3CA | 0.648722055 | 3.886448004 | 0.000241564 | 0.006765746 | protein phosphatase 3 catalytic subunit alpha | PPP3CA.PPP3R1 |
| PRTN3 | 1.157976164 | 3.772549339 | 0.000352163 | 0.00913468 | proteinase 3 | |
| COLEC12 | -0.748316044 | -3.758701358 | 0.000368529 | 0.00918431 | collectin subfamily member 12 | |
| CTSV | -0.737192857 | -3.736622616 | 0.000396135 | 0.009499762 | cathepsin V | |
| BCAN | -0.624746995 | -3.714762031 | 0.000425409 | 0.010012867 | brevican | |
| FYN | -1.086478202 | -3.690449924 | 0.000460391 | 0.010639219 | FYN proto-oncogene, Src family tyrosine kinase | |

| | | | | | | |
|---|---|---|---|---|---|---|
| ITGA2B | -0.805740518 | -3.682073075 | 0.000473068 | 0.010736956 | integrin subunit alpha 2b | ITGA2B.ITGB3 |
| IGHA1 | 0.664989929 | 3.61280457 | 0.000591436 | 0.012528588 | immunoglobulin heavy constant alpha 1 | IGHA1.IGHA2 |
| LCN2 | 1.145530501 | 3.616246758 | 0.000584941 | 0.012528588 | lipocalin 2 | |
| KYNU | 0.986816086 | 3.599836103 | 0.000616527 | 0.012846003 | kynureninase | |
| IL36A | -1.723984034 | -3.561804217 | 0.000696085 | 0.014043241 | interleukin 36 alpha | |
| FCGR3B | 0.995678631 | 3.467970518 | 0.000936189 | 0.017759653 | Fc fragment of IgG receptor IIIb | |
| CRP | 2.015314074 | 3.462928471 | 0.000951096 | 0.017777102 | C-reactive protein | |
| MRC1 | 0.633109586 | 3.456230272 | 0.000971247 | 0.017890649 | mannose receptor C-type 1 | |
| RETN | 0.70260533 | 3.448366893 | 0.000995419 | 0.018040681 | resistin | |
| KLK8 | -0.675000668 | -3.429183567 | 0.001056803 | 0.018180859 | kallikrein related peptidase 8 | |
| MRC2 | -0.676330115 | -3.428661029 | 0.001058524 | 0.018180859 | mannose receptor C type 2 | |
| FGR | 1.031226964 | 3.403270748 | 0.001145424 | 0.019402124 | FGR proto-oncogene, Src family tyrosine kinase | |
| MIA | -0.608824356 | -3.393689676 | 0.001179936 | 0.019476614 | melanoma inhibitory activity | |
| OMD | -0.772947597 | -3.378591114 | 0.001236325 | 0.020145765 | osteomodulin | |
| DLL4 | 0.622838128 | 3.349363268 | 0.001352793 | 0.021492495 | delta like canonical Notch ligand 4 | |
| ARG1 | 0.937212532 | 3.200335815 | 0.002125637 | 0.030018716 | arginase 1 | |
| F9 | 0.694989244 | 3.185456224 | 0.002222259 | 0.031038371 | coagulation factor IX | |
| ACAN | -0.789557187 | -3.110973501 | 0.002770931 | 0.035937276 | aggrecan | |
| IL6 | 0.875184946 | 3.088483302 | 0.002960021 | 0.038001884 | interleukin 6 | |
| CCL16 | 0.80302904 | 3.039429964 | 0.00341499 | 0.042861872 | C-C motif chemokine ligand 16 | |
| F2 | -0.586859929 | -3.020469738 | 0.003607689 | 0.043670219 | coagulation factor II, thrombin | |
| IMPDH1 | 1.092833534 | 3.003950686 | 0.003783778 | 0.043719831 | inosine monophosphate dehydrogenase 1 | |
| CD163 | 0.60137612 | 2.993530975 | 0.003898925 | 0.044497858 | CD163 molecule | |
| MST1 | -0.89971528 | -2.921621891 | 0.004786733 | 0.051558793 | macrophage stimulating 1 | |
| ANXA1 | 0.770844072 | 2.831994123 | 0.006154824 | 0.06359985 | annexin A1 | |
| BPI | 1.338630343 | 2.76763689 | 0.00735032 | 0.073561072 | bactericidal/permeability-increasing protein | |
| IL1RL1 | 0.98512885 | 2.759642807 | 0.007512854 | 0.074600291 | interleukin 1 receptor like 1 | |
| HNRNPA2B1 | 0.796750709 | 2.742829334 | 0.007865514 | 0.076900526 | heterogeneous nuclear ribonucleoprotein A2/B1 | |
| ADRBK1 | 0.629098886 | 2.706686303 | 0.00867551 | 0.080485937 | NA | |
| CDON | -0.622995579 | -2.709389463 | 0.008612387 | 0.080485937 | cell adhesion associated, oncogene regulated | |
| TNFAIP6 | 0.606792963 | 2.710062752 | 0.008596731 | 0.080485937 | TNF alpha induced protein 6 | |
| TFF1 | 0.736084344 | 2.67195038 | 0.009525155 | 0.086474801 | trefoil factor 1 | |
| XRCC6 | 0.597138465 | 2.67226078 | 0.009517238 | 0.086474801 | X-ray repair cross complementing 6 | |
| CA6 | -1.039389931 | -2.636474725 | 0.010470638 | 0.091780554 | carbonic anhydrase 6 | |
| EPO | 0.657346475 | 2.630858116 | 0.01062793 | 0.09189183 | erythropoietin | |
| CA3 | -0.679266429 | -2.601498958 | 0.011485664 | 0.097975029 | carbonic anhydrase 3 | |
| HGF | 0.60288153 | 2.590043224 | 0.011837059 | 0.09897962 | hepatocyte growth factor | |
| MMP8 | 1.064276273 | 2.448053123 | 0.017076939 | 0.130704429 | matrix metallopeptidase 8 | |
| MMP9 | 0.664360023 | 2.448485006 | 0.017058253 | 0.130704429 | matrix metallopeptidase 9 | |

| | | | | | | |
|---|---|---|---|---|---|---|
| ICOSLG | 0.60206072 | 2.432021248 | 0.017783716 | 0.134542282 | inducible T-cell costimulator ligand | |
| COL8A1 | 0.672616053 | 2.416875883 | 0.018475385 | 0.135733565 | collagen type VIII alpha 1 chain | |
| SFTPD | 1.032516415 | 2.415991846 | 0.018516493 | 0.135733565 | surfactant protein D | |
| MAPK1 | 0.686684336 | 2.40106143 | 0.019223227 | 0.13756474 | mitogen-activated protein kinase 1 | |
| PPIF | -0.799374275 | -2.367748533 | 0.02088772 | 0.145072634 | peptidylprolyl isomerase F | |
| AKR1A1 | 0.605561556 | 2.318367131 | 0.023591732 | 0.154093001 | aldo-keto reductase family 1 member A1 | |
| LTA4H | -1.444148219 | -2.309253013 | 0.024123478 | 0.154824019 | leukotriene A4 hydrolase | |
| PLA2G2A | 0.990721527 | 2.27157691 | 0.026436103 | 0.163903839 | phospholipase A2 group IIA | |
| MDK | -0.806953514 | -2.222574853 | 0.029735905 | 0.174287384 | midkine | |
| CD177 | 0.689873471 | 2.186543509 | 0.032388365 | 0.179898302 | CD177 molecule | |
| PTPN11 | -0.606073471 | -2.169812105 | 0.033689052 | 0.182173461 | protein tyrosine phosphatase, non-receptor type 11 | |
| HSP90AA1 | 0.605987126 | 2.16499627 | 0.034071805 | 0.182722633 | heat shock protein 90 alpha family class A member 1 | HSP90AA1.HSP90AB1 |

*: If a gene symbol is mapped to multiple annotated SOMAscan analytes (protein or protein complexes), the information is shown.

**Table S3: Proteins whose concentration was significantly altered in the MAS disease component**

| Gene Symbol ID | Coefficients | t | P.Value | FDR | full_name | Prt/Prt Complex mapped* |
|---|---|---|---|---|---|---|
| FTH1 | 3.399631014 | 8.478480327 | 4.19E-12 | 5.32E-09 | ferritin heavy chain 1 | FTH1.FTL |
| HSPA1A | 1.303989558 | 6.287296851 | 3.10E-08 | 9.86E-06 | heat shock protein family A (Hsp70) member 1A | |
| MAP2K1 | 0.615474776 | 6.312923966 | 2.80E-08 | 9.86E-06 | mitogen-activated protein kinase kinase 1 | |
| SERPINA4 | -0.937763489 | -6.405068942 | 1.94E-08 | 9.86E-06 | serpin family A member 4 | |
| YES1 | 0.911991643 | 5.891712499 | 1.49E-07 | 3.80E-05 | YES proto-oncogene 1, Src family tyrosine kinase | |
| ENO1 | 1.274696374 | 5.471017697 | 7.71E-07 | 0.000139934 | enolase 1 | |
| MAPK14 | 1.34176103 | 5.359046795 | 1.19E-06 | 0.000188346 | mitogen-activated protein kinase 14 | |
| HSP90AA1 | 1.541717073 | 5.089897471 | 3.30E-06 | 0.000465573 | heat shock protein 90 alpha family class A member 1 | HSP90AA1.HSP90AB1 |
| AFM | -0.811793112 | -4.923343068 | 6.15E-06 | 0.000723785 | afamin | |
| HSPA8 | 1.193320113 | 4.805976323 | 9.49E-06 | 0.000928127 | heat shock protein family A (Hsp70) member 8 | |
| PLA2G2A | 2.537238761 | 4.812063239 | 9.28E-06 | 0.000928127 | phospholipase A2 group IIA | |
| HSP90AB1 | 1.067570601 | 4.780609148 | 1.04E-05 | 0.000946137 | heat shock protein 90 alpha family class B member 1 | |
| HIST1H3A | 1.553006523 | 4.545101983 | 2.45E-05 | 0.001633146 | histone cluster 1 H3 family member a | |
| LBP | 0.791793981 | 4.538426099 | 2.51E-05 | 0.001633146 | lipopolysaccharide binding protein | |
| PRKCZ | 0.804760109 | 4.554154518 | 2.38E-05 | 0.001633146 | protein kinase C zeta | |
| THBS2 | 1.251949261 | 4.532292527 | 2.57E-05 | 0.001633146 | thrombospondin 2 | |
| CA6 | -1.839730651 | -4.434859636 | 3.64E-05 | 0.002203595 | carbonic anhydrase 6 | |
| NAMPT | 1.600166221 | 4.278475234 | 6.32E-05 | 0.003493788 | nicotinamide phosphoribosyltransferase | |
| DCTPP1 | 1.195168831 | 4.210757523 | 8.01E-05 | 0.004205713 | dCTP pyrophosphatase 1 | |
| MPO | 0.954850612 | 4.201313817 | 8.27E-05 | 0.004205713 | myeloperoxidase | |
| FCN3 | 0.822268491 | 4.159890378 | 9.55E-05 | 0.00449439 | ficolin 3 | |
| NAGK | 1.659404418 | 4.087772132 | 0.000122335 | 0.005361643 | N-acetylglucosamine kinase | |
| MAP2K4 | -1.379796553 | -4.003671051 | 0.000162896 | 0.006678716 | mitogen-activated protein kinase kinase 4 | |
| GDI2 | 0.918223052 | 3.895542819 | 0.000234342 | 0.009006606 | GDP dissociation inhibitor 2 | |
| MCL1 | 0.798717224 | 3.873765594 | 0.000251993 | 0.009006606 | MCL1, BCL2 family apoptosis regulator | |
| MMP17 | 0.595401711 | 3.865455598 | 0.000259059 | 0.009006606 | matrix metallopeptidase 17 | |
| LDHB | 0.707544413 | 3.84726616 | 0.000275195 | 0.009204546 | lactate dehydrogenase B | |
| STAT3 | 0.9387298 | 3.816673705 | 0.000304529 | 0.00992451 | signal transducer and activator of transcription 3 | |
| AKR1A1 | 1.247129117 | 3.801542652 | 0.000320122 | 0.01017188 | aldo-keto reductase family 1 member A1 | |
| POR | 1.159717431 | 3.79144723 | 0.000330948 | 0.010259396 | cytochrome p450 oxidoreductase | |
| GPI | 1.25843132 | 3.776058904 | 0.000348127 | 0.010330923 | glucose-6-phosphate isomerase | |
| FGG | 0.678108073 | 3.705127656 | 0.000438957 | 0.011870509 | fibrinogen gamma chain | |
| MDH1 | 0.880344575 | 3.714716428 | 0.000425472 | 0.011870509 | malate dehydrogenase 1 | |
| XPNPEP1 | 1.387740104 | 3.707461407 | 0.000435638 | 0.011870509 | X-prolyl aminopeptidase 1 | |
| CD27 | -0.614582841 | -3.638956661 | 0.000543764 | 0.014094009 | CD27 molecule | |
| HGFAC | -1.085257764 | -3.617229109 | 0.0005831 | 0.014531774 | HGF activator | |
| CAPG | 0.827240288 | 3.59765475 | 0.000620846 | 0.014574341 | capping actin protein, gelsolin like | |
| RBM39 | 0.781140644 | 3.57320891 | 0.000671257 | 0.014967852 | RNA binding motif protein 39 | |
| ICOSLG | 1.09429432 | 3.523717385 | 0.000785479 | 0.01692109 | inducible T-cell costimulator ligand | |
| APOM | -0.756921528 | -3.485757829 | 0.000885352 | 0.018447245 | apolipoprotein M | |
| YWHAB | 0.880605704 | 3.4468683 | 0.00100009 | 0.019861167 | tyrosine 3-monooxygenase/tryptophan 5-monooxygenase activation protein beta | YWHAB, YWHAB.YWHAE.YWHAG.YWHAH.YWHAQ.YWHAZ.SFN |
| CTSV | -0.737519067 | -3.42738789 | 0.001062728 | 0.020465569 | cathepsin V | |
| PTPN11 | 1.103376202 | 3.400491644 | 0.001155335 | 0.021621756 | protein tyrosine phosphatase, non-receptor type 11 | |
| HTRA2 | 0.690483266 | 3.313543474 | 0.00150966 | 0.025247071 | HtrA serine peptidase 2 | |
| HRG | -0.672050266 | -3.304348924 | 0.001552607 | 0.025628101 | histidine rich glycoprotein | |
| PTPN6 | 1.261684265 | 3.290160501 | 0.001621146 | 0.026081984 | protein tyrosine phosphatase, non-receptor type 6 | |
| YWHAZ | 0.819023932 | 3.259602278 | 0.001778554 | 0.027907926 | tyrosine 3-monooxygenase/tryptophan 5-monooxygenase activation protein zeta | |
| IL1RL1 | 1.406019984 | 3.240846423 | 0.001882171 | 0.028479036 | interleukin 1 receptor like 1 | |
| CLEC1B | -0.597932304 | -3.115655415 | 0.002733014 | 0.037592137 | C-type lectin domain family 1 member B | |
| PRTN3 | 1.030316413 | 3.120927359 | 0.002690899 | 0.037592137 | proteinase 3 | |
| ASAH2 | -0.792568667 | -3.067844064 | 0.003144084 | 0.041197229 | N-acylsphingosine amidohydrolase 2 | |
| MAPK3 | 0.725255462 | 3.069047525 | 0.003133065 | 0.041197229 | mitogen-activated protein kinase 3 | |
| MRC1 | 0.633877747 | 3.048621504 | 0.003325063 | 0.042688439 | mannose receptor C-type 1 | |
| ADRBK1 | 0.817521788 | 3.036283396 | 0.003446291 | 0.042943494 | NA | |
| IGFBP2 | 1.035643888 | 3.04031507 | 0.003406233 | 0.042943494 | insulin like growth factor binding protein 2 | |
| FAP | -0.599323521 | -2.985943578 | 0.003984812 | 0.048235197 | fibroblast activation protein alpha | |

| | | | | | | |
|---|---|---|---|---|---|---|
| THPO | 0.667916412 | 2.972485846 | 0.004141486 | 0.049658767 | thrombopoietin | |
| BMP10 | 0.675041353 | 2.921310717 | 0.004790952 | 0.0538632 | bone morphogenetic protein 10 | |
| CD36 | -0.726836342 | -2.907153597 | 0.004986602 | 0.0538632 | CD36 molecule | |
| IL18 | 3.464802204 | 2.9031643 | 0.005043053 | 0.0538632 | interleukin 18 | |
| PYY | -0.624946946 | -2.906048671 | 0.005002178 | 0.0538632 | peptide YY | |
| FCER2 | 0.713454419 | 2.822041425 | 0.006327167 | 0.060013651 | Fc fragment of IgE receptor II | |
| PRDX1 | 0.702448006 | 2.778690918 | 0.007130885 | 0.064738253 | peroxiredoxin 1 | |
| ITGA1 | 0.792001293 | 2.763270889 | 0.007438684 | 0.066067905 | integrin subunit alpha 1 | ITGA1.ITGB1 |
| HAMP | 1.610634376 | 2.740690335 | 0.007911451 | 0.067486272 | hepcidin antimicrobial peptide | |
| VWF | 0.877958445 | 2.713053642 | 0.008527494 | 0.071777781 | von Willebrand factor | |
| STAT1 | 1.041445649 | 2.688571327 | 0.009109554 | 0.075183399 | signal transducer and activator of transcription 1 | |
| PDGFB | -0.84472126 | -2.631373377 | 0.010613411 | 0.082367421 | platelet derived growth factor subunit B | |
| PGAM1 | 1.379972744 | 2.63484296 | 0.010516115 | 0.082367421 | phosphoglycerate mutase 1 | |
| RAC1 | 0.700934055 | 2.630853706 | 0.010628054 | 0.082367421 | Rac family small GTPase 1 | |
| PDGFA | -0.591292855 | -2.618094025 | 0.010993411 | 0.084682579 | platelet derived growth factor subunit A | |
| UBE2I | 0.643534564 | 2.584284663 | 0.012017357 | 0.088364403 | ubiquitin conjugating enzyme E2 I | |
| TPI1 | 0.664833687 | 2.547220745 | 0.013238638 | 0.092963031 | triosephosphate isomerase 1 | |
| SHC1 | 0.61211794 | 2.542241572 | 0.013410993 | 0.093163162 | SHC adaptor protein 1 | |
| TKT | 0.591631136 | 2.513994728 | 0.014427732 | 0.097331052 | transketolase | |
| F2 | -0.642474418 | -2.496164139 | 0.015104839 | 0.0988685 | coagulation factor II, thrombin | |
| PI3 | 0.755231593 | 2.492526607 | 0.015246441 | 0.0988685 | peptidase inhibitor 3 | |
| PRSS2 | 0.721001898 | 2.493595267 | 0.015204716 | 0.0988685 | protease, serine 2 | |
| CAT | 0.685412674 | 2.472067566 | 0.016065372 | 0.101587501 | catalase | |
| IGHM | -0.641461834 | -2.460519006 | 0.01654491 | 0.102348016 | immunoglobulin heavy constant mu | IGHM.IGJ.IGK..IGL. |
| SAA1 | 2.21597527 | 2.460357869 | 0.016551691 | 0.102348016 | serum amyloid A1 | |
| CXCL13 | 0.860900442 | 2.452770014 | 0.016873848 | 0.10302182 | C-X-C motif chemokine ligand 13 | |
| TNFRSF1B | 0.699905341 | 2.445227046 | 0.017199668 | 0.10311688 | TNF receptor superfamily member 1B | |
| ADSL | 0.652828881 | 2.430864752 | 0.017835701 | 0.104949892 | adenylosuccinate lyase | |
| NTN1 | 1.029074333 | 2.416432513 | 0.018495992 | 0.108333666 | netrin 1 | |
| RET | -0.596834722 | -2.399213415 | 0.01931236 | 0.110690068 | ret proto-oncogene | |
| BAD | 0.591212373 | 2.380230589 | 0.020249553 | 0.112953495 | BCL2 associated agonist of cell death | |
| S100A12 | 1.127968291 | 2.378219535 | 0.02035118 | 0.112953495 | S100 calcium binding protein A12 | |
| S100A9 | 0.651480136 | 2.371240875 | 0.020707385 | 0.113825083 | S100 calcium binding protein A9 | |
| TPT1 | 0.684222325 | 2.366045806 | 0.020976151 | 0.114423553 | tumor protein, translationally-controlled 1 | |
| GNLY | 0.734217793 | 2.351913751 | 0.021723075 | 0.116498009 | granulysin | |
| FCGR3B | 0.91875608 | 2.330275057 | 0.022912679 | 0.119352523 | Fc fragment of IgG receptor IIIb | |
| CDON | -0.668255305 | -2.265795398 | 0.026807816 | 0.133618566 | cell adhesion associated, oncogene regulated | |
| LEP | -0.691993914 | -2.206500176 | 0.030894688 | 0.144397064 | leptin | |
| HIST3H2A | 1.103189207 | 2.195070309 | 0.031742591 | 0.14657682 | histone cluster 3 H2A | |
| LAG3 | 0.588793562 | 2.181755092 | 0.032756041 | 0.149221964 | lymphocyte activating 3 | |
| SUMO3 | 0.634822297 | 2.176025308 | 0.033200801 | 0.149755012 | small ubiquitin-like modifier 3 | |
| HIST2H2BE | 0.758951347 | 2.171555168 | 0.033551447 | 0.15068512 | histone cluster 2 H2B family member e | |
| LGALS9 | 0.60516485 | 2.10918635 | 0.038792385 | 0.163441503 | galectin 9 | |
| IL1R1 | -1.612387882 | -2.097649429 | 0.039836242 | 0.166552184 | interleukin 1 receptor type 1 | |
| MDK | 0.902523744 | 2.082286681 | 0.041264059 | 0.170835893 | midkine | |
| CRP | 1.246753165 | 2.063024243 | 0.043116812 | 0.17677893 | C-reactive protein | |
| PRKACA | 0.837879231 | 2.05864277 | 0.043548149 | 0.177402875 | protein kinase cAMP-activated catalytic subunit alpha | |
| CXCL9 | 0.973464496 | 2.052831573 | 0.044125985 | 0.177481413 | C-X-C motif chemokine ligand 9 | |
| LCN2 | 0.813689672 | 2.053995424 | 0.04400973 | 0.177481413 | lipocalin 2 | |

*: If a gene symbol is mapped to multiple annotated SOMAscan analytes (protein or protein complexes), the information is shown.

#### Table S4: Proteins whose concentration was significantly altered in the sJIA-PAP disease component

| Gene Symbol ID | Coefficients | t | P.Value | FDR | full_name | Prt/Prt Complex mapped* |
|---|---|---|---|---|---|---|
| ICAM5 | 0.846443494 | 5.139654079 | 2.07E-06 | 0.000916677 | intercellular adhesion molecule 5 | |
| POR | 1.082182934 | 5.129217848 | 2.16E-06 | 0.000916677 | cytochrome p450 oxidoreductase | |
| MMP7 | 0.927518201 | 5.102130708 | 2.41E-06 | 0.000916677 | matrix metallopeptidase 7 | |
| CCL11 | 0.708451121 | 4.990188794 | 3.73E-06 | 0.000916677 | C-C motif chemokine ligand 11 | |
| ANG | 0.721309076 | 4.956708219 | 4.25E-06 | 0.000916677 | angiogenin | |
| GDF15 | 0.775165598 | 4.951867243 | 4.33E-06 | 0.000916677 | growth differentiation factor 15 | |
| CAPN1 | 0.62832412 | 4.908995558 | 5.11E-06 | 0.000927715 | calpain 1 | CAPN1.CAPNS1 |
| LDHB | 0.587787775 | 4.681692829 | 1.22E-05 | 0.001936829 | lactate dehydrogenase B | |
| S100A12 | -1.664300731 | -4.588267529 | 1.73E-05 | 0.002447341 | S100 calcium binding protein A12 | |
| APOM | 0.711000075 | 4.432038779 | 3.10E-05 | 0.003523465 | apolipoprotein M | |
| REN | 0.915497396 | 4.259051747 | 5.82E-05 | 0.005686816 | renin | |
| FLRT3 | 0.62081125 | 4.173446337 | 7.91E-05 | 0.007179121 | fibronectin leucine rich transmembrane protein 3 | |
| FUT5 | -0.689397814 | -4.00203719 | 0.000144778 | 0.012267512 | fucosyltransferase 5 | |
| CCL15 | 0.760877528 | 3.951247772 | 0.000172724 | 0.013669992 | C-C motif chemokine ligand 15 | |
| RET | -0.720960447 | -3.673599667 | 0.000443026 | 0.024481977 | ret proto-oncogene | |
| HAMP | -1.899087025 | -3.632496994 | 0.000507583 | 0.026880747 | hepcidin antimicrobial peptide | |
| NAGK | 0.828559321 | 3.586803995 | 0.00058982 | 0.029986474 | N-acetylglucosamine kinase | |
| CCL7 | 0.927730591 | 3.541589425 | 0.00068354 | 0.033414606 | C-C motif chemokine ligand 7 | |
| CCDC80 | -0.593197029 | -3.438295419 | 0.000953305 | 0.04060835 | coiled-coil domain containing 80 | |
| F5 | 0.606823905 | 3.436593001 | 0.000958498 | 0.04060835 | coagulation factor V | |
| CCL17 | 0.971315296 | 3.276944482 | 0.001583552 | 0.062896703 | C-C motif chemokine ligand 17 | |
| CCL25 | 0.883134117 | 3.234396174 | 0.00180574 | 0.069253778 | C-C motif chemokine ligand 25 | |
| KYNU | 0.927687027 | 3.226057059 | 0.001852579 | 0.069253778 | kynureninase | |
| IL18 | 2.269773632 | 3.05371015 | 0.003115718 | 0.099452085 | interleukin 18 | |
| MMP1 | -0.91980654 | -3.016945868 | 0.003472961 | 0.107661789 | matrix metallopeptidase 1 | |
| ADIPOQ | 0.607212126 | 2.930433231 | 0.004468754 | 0.126217485 | adiponectin, C1Q and collagen domain containing | |

*: If a gene symbol is mapped to multiple annotated SOMAscan analytes (protein or protein complexes), the information is shown.

### Table S5: Gene Ontology Enrichment for different disease components

| Comparison | set.name | original.size | filtered.size | relevant.protein.size | relevant.proteins | pval | padj |
|---|---|---|---|---|---|---|---|
| Active sJIA vs. Healthy control | GO_MYELOID_LEUKOCYTE_MEDIATED_IMMUNITY | 555 | 147 | 25 | LILRB2,MAPK14,F2,FCGR3B,FGR,FTH1,XRCC6,GPI,HSP90AA1,IL6,IMPDH1,ARG1,LCN2,LTA4H,MMP8,MMP9,PKM,MAPK1,PRTN3,RETN,CD177,S100A9,S100A12,BPI,TNFAIP6 | 4.16E-06 | 0.008897 |
| Active sJIA vs. Healthy control | GO_COLLAGEN_METABOLIC_PROCESS | 113 | 33 | 10 | F2,FAP,IL6,ARG1,MMP1,MMP3,MMP8,MMP9,PRTN3,MRC2 | 3.25E-05 | 0.023738 |
| Active sJIA vs. Healthy control | GO_CELL_ACTIVATION_INVOLVED_IN_IMMUNE_RESPONS | 720 | 187 | 27 | LILRB2,MAPK14,FCGR3B,FGR,FTH1,XRCC6,GPI,ANXA1,HSP90AA1,IL6,IL18,IMPDH1,ARG1,LCN2,LTA4H,MDK,MMP8,MMP9,PKM,MAPK1,PRTN3,RETN,CD177,S100A9,S100A12,BPI,TNFAIP6 | 4.01E-05 | 0.023738 |
| Active sJIA vs. Healthy control | GO_MYELOID_LEUKOCYTE_ACTIVATION | 662 | 188 | 27 | LILRB2,MAPK14,FCGR3B,FGR,FTH1,XRCC6,GPI,HSP90AA1,IL6,IL18,IMPDH1,ARG1,LCN2,LTA4H,MMP8,MMP9,PKM,PLA2G2A,MAPK1,PRTN3,RETN,CD177,S100A9,S100A12,BPI,TNFAIP6,IL1RL1 | 4.44E-05 | 0.023738 |
| Active sJIA vs. Healthy control | GO_EXOCYTOSIS | 914 | 203 | 27 | LILRB2,CD109,MAPK14,FCGR3B,FGR,FTH1,XRCC6,GPI,ANXA1,HGF,HSP90AA1,IMPDH1,ITGA2B,ARG1,LCN2,LTA4H,MMP8,MMP9,PKM,MAPK1,PRTN3,RETN,CD177,S100A9,S100A12,BPI,TNFAIP6 | 0.000184 | 0.069897 |
| Active sJIA vs. Healthy control | GO_CELL_ACTIVATION | 1475 | 374 | 41 | LILRB2,MAPK14,EPO,F2,FCGR3B,FGR,ICOSLG,FTH1,FYN,XRCC6,GPI,ANXA1,HSP90AA1,IGHA1,IL6,IL18,IMPDH1,ITGA2B,ARG1,LCN2,LTA4H,MDK,MMP8,MMP9,PKM,PLA2G2A,DLL4,PPP3CA,MAPK1,MAPK3,PRTN3,RETN,CD177,PTPN11,S100A9,S100A12,SAA1,SFTPD,BPI,TNFAIP6,IL1RL1 | 0.000196 | 0.069897 |
| Active sJIA vs. Healthy control | GO_MONOSACCHARIDE_CATABOLIC_PROCESS | 66 | 16 | 6 | AKR1A1,FUT5,GAPDH,GPI,HK2,PKM | 0.000388 | 0.084411 |
| Active sJIA vs. Healthy control | GO_RESPONSE_TO_BACTERIUM | 743 | 178 | 24 | LILRB2,CRP,MAPK14,EPO,F2,FGR,IL36A,GPC3,APOB,IGHA1,IL6,IL18,ARG1,LCN2,MRC1,PLA2G2A,MAPK1,MAPK3,S100A9,S100A12,SFTPD,BPI,CA3,COLEC12 | 0.000388 | 0.084411 |
| Active sJIA vs. Healthy control | GO_COLLAGEN_CATABOLIC_PROCESS | 47 | 22 | 7 | FAP,MMP1,MMP3,MMP8,MMP9,PRTN3,MRC2 | 0.000394 | 0.084411 |
| Active sJIA vs. Healthy control | GO_MACROPHAGE_CHEMOTAXIS | 42 | 22 | 7 | MAPK14,MDK,MST1,MAPK1,MAPK3,SAA1,SFTPD | 0.000394 | 0.084411 |
| Active sJIA vs. Healthy control | GO_LEUKOCYTE_MEDIATED_IMMUNITY | 885 | 225 | 28 | LILRB2,CRP,MAPK14,F2,FCGR3B,FGR,FTH1,XRCC6,GPI,HSP90AA1,IGHA1,IL6,IL18,IMPDH1,ARG1,LCN2,LTA4H,MMP8,MMP9,PKM,MAPK1,PRTN3,RETN,CD177,S100A9,S100A12,BPI,TNFAIP6 | 0.000445 | 0.086609 |
| Active sJIA vs. Healthy control | GO_ORGANIC_ACID_METABOLIC_PROCESS | 1183 | 118 | 18 | AKR1A1,CKB,CKM,MAPK14,ACAN,GAPDH,GPI,ANXA1,HGF,HK2,ARG1,LTA4H,OMD,PKM,NAGK,MAPK3,BCAN,KYNU | 0.000549 | 0.097866 |
| Active sJIA vs. Healthy control | GO_SECRETION | 1618 | 331 | 36 | LILRB2,CD109,MAPK14,GRK2,FCGR3B,FGR,FTH1,XRCC6,GAPDH,GPI,ANXA1,HGF,HK2,HSP90AA1,IL6,IMPDH1,ITGA2B,ARG1,LCN2,LTA4H,MMP8,MMP9,PKM,PLA2G2A,PPP3CA,MAPK1,PRTN3,RETN,CD177,PTPN11,S100A9,S100A12,SAA1,BPI,TNFAIP6,IL1RL1 | 0.000789 | 0.12557 |
| Active sJIA vs. Healthy control | GO_CELLULAR_RESPONSE_TO_CHEMICAL_STRESS | 360 | 83 | 14 | PPIF,EPO,FYN,XRCC6,ANXA1,HGF,IL6,ARG1,LCN2,MCL1,MMP3,MMP9,MAPK1,MAPK3 | 0.000895 | 0.12557 |
| Active sJIA vs. Healthy control | GO_MONOCARBOXYLIC_ACID_METABOLIC_PROCESS | 672 | 74 | 13 | AKR1A1,CKB,CKM,MAPK14,GAPDH,GPI,ANXA1,HK2,LTA4H,PKM,MAPK3,BCAN,KYNU | 0.000934 | 0.12557 |
| Active sJIA vs. Healthy control | GO_MACROPHAGE_MIGRATION | 57 | 25 | 7 | MAPK14,MDK,MST1,MAPK1,MAPK3,SAA1,SFTPD | 0.000939 | 0.12557 |
| Active sJIA vs. Healthy control | GO_ANTIMICROBIAL_HUMORAL_RESPONSE | 137 | 49 | 10 | F2,GAPDH,IGHA1,LCN2,PLA2G2A,PRTN3,S100A9,S100A12,SFTPD,BPI | 0.001158 | 0.137721 |
| Active sJIA vs. Healthy control | GO_SMALL_MOLECULE_CATABOLIC_PROCESS | 452 | 49 | 10 | AKR1A1,FUT5,GAPDH,GPI,GSK3A,HK2,ARG1,PKM,NAGK,KYNU | 0.001158 | 0.137721 |
| MAS vs. active sJIA | GO_ANTIMICROBIAL_HUMORAL_RESPONSE | 137 | 49 | 14 | CXCL13,GNLY,F2,HRG,IGHM,LCN2,CXCL9,PI3,PLA2G2A,PRSS2,PRTN3,S100A9,S100A12,H2BC21 | 1.86E-05 | 0.044402 |
| MAS vs. active sJIA | GO_CELL_ACTIVATION | 1475 | 374 | 50 | NAMPT,MAPK14,F2,FCGR3B,FGG,ICOSLG,FTH1,GDI2,GPI,HRG,HSPA1A,HSPA8,HSP90AA1,HSP90AB1,IGFBP2,IGHM,IL18,LAG3,LBP,LCN2,LEP,LGALS9,MDK,MPO,PRDX1,CLEC1B,PDGFA,PDGFB,PGAM1,PLA2G2A,PRKCZ,MAPK3,PRSS2,PRTN3,BAD,PTPN6,PTPN11,RAC1,S100A9,S100A12,SAA1,STAT3,TNFRSF1B,VWF,YES1,YWHAZ,CAT,IL1RL1,CD27,CD36 | 3.73E-05 | 0.04441 |
| MAS vs. active sJIA | GO_COAGULATION | 349 | 95 | 19 | F2,FAP,FGG,HGFAC,HRG,CLEC1B,PDGFA,PDGFB,PRKACA,MAPK3,PRTN3,PTPN6,PTPN11,RAC1,SAA1,VWF,YWHAZ,H3C1,CD36 | 0.000141 | 0.098608 |
| MAS vs. active sJIA | GO_IMMUNE_EFFECTOR_PROCESS | 1292 | 300 | 41 | CRP,MAPK14,F2,FCER2,FCGR3B,FTH1,GDI2,GPI,HSPA1A,HSPA8,HSP90AA1,HSP90AB1,IGHM,IL1R1,IL18,LAG3,LBP,LCN2,LEP,LGALS9,MDK,CXCL9,MPO,PRDX1,PGAM1,PRKCZ,MAPK3,PRSS2,PRTN3,PTPN6,RAC1,S100A9,S100A12,STAT1,STAT3,TNFRSF1B,YES1,CAT,FCN3,CD27,CD36 | 0.00019 | 0.098608 |
| MAS vs. active sJIA | GO_PLATELET_ACTIVATION | 158 | 46 | 12 | F2,FGG,HRG,CLEC1B,PDGFA,PDGFB,MAPK3,PTPN6,PTPN11,SAA1,VWF,YWHAZ | 0.000207 | 0.098608 |
| MAS vs. active sJIA | GO_LEUKOCYTE_MEDIATED_IMMUNITY | 885 | 225 | 33 | CRP,MAPK14,F2,FCER2,FCGR3B,FTH1,GDI2,GPI,HSPA1A,HSPA8,HSP90AA1,HSP90AB1,IGHM,IL1R1,IL18,LAG3,LCN2,LEP,LGALS9,MPO,PRDX1,PGAM1,PRKCZ,PRSS2,PRTN3,PTPN6,RAC1,S100A9,S100A12,TNFRSF1B,CAT,CD27,CD36 | 0.00028 | 0.111357 |
| MAS vs. active sJIA | GO_DEFENSE_RESPONSE_TO_BACTERIUM | 341 | 81 | 16 | CXCL13,GNLY,CRP,F2,IGHM,LBP,LCN2,MPO,PI3,PLA2G2A,HAMP,S100A9,S100A12,SHC1,H2BC21,CD36 | 0.000593 | 0.185039 |
| MAS vs. active sJIA | GO_REGULATION_OF_BODY_FLUID_LEVELS | 513 | 115 | 20 | F2,FAP,FGG,GPI,HGFAC,HRG,CLEC1B,PDGFA,PDGFB,PRKACA,MAPK3,PRTN3,PTPN6,PTPN11,RAC1,SAA1,VWF,YWHAZ,H3C1,CD36 | 0.000685 | 0.185039 |
| MAS vs. active sJIA | GO_HUMORAL_IMMUNE_RESPONSE | 370 | 107 | 19 | CXCL13,GNLY,CRP,F2,FCER2,GPI,HRG,IGHM,LCN2,CXCL9,PI3,PLA2G2A,PRSS2,PRTN3,PTPN6,S100A9,S100A12,H2BC21,FCN3 | 0.000726 | 0.185039 |
| MAS vs. active sJIA | GO_MONOSACCHARIDE_BIOSYNTHETIC_PROCESS | 100 | 26 | 8 | AKR1A1,ENO1,GPI,LEP,MDH1,PGAM1,TKT,TPI1 | 0.000777 | 0.185039 |
| MAS vs. active sJIA | GO_OXIDATION_REDUCTION_PROCESS | 1014 | 84 | 16 | AKR1A1,MAPK14,ADSL,ENO1,FTH1,GPI,LDHB,LEP,MDH1,MPO,PRDX1,PGAM1,POR,TKT,TPI1,CAT | 0.000909 | 0.190158 |
| MAS vs. active sJIA | GO_REGULATION_OF_RESPONSE_TO_STRESS | 1538 | 332 | 42 | NAMPT,MAPK14,ENO1,F2,FAP,FGG,BMP10,HTRA2,HRG,HSPA1A,HSPA8,HSP90AA1,HSP90AB1,IL1R1,IL18,LAG3,LBP,LEP,LGALS9,MCL1,MDK,PRDX1,PDGFA,PDGFB,PLA2G2A,PRKACA,MAPK3,MAP2K1,BAD,PTPN6,PTPN11,S100A9,S100A12,SAA1,MAP2K4,STAT1,STAT3,TNFRSF1B,TPT1,IL1RL1,CD27,CD36 | 0.000958 | 0.190158 |
| sJIA-PAP vs all sJIA | GO_MONOCYTE_CHEMOTAXIS | 66 | 44 | 6 | S100A12,CCL7,CCL11,CCL15,CCL17,CCL25 | 0.000173 | 0.16479 |
| sJIA-PAP vs all sJIA | GO_LYMPHOCYTE_MIGRATION | 116 | 52 | 6 | RET,CCL7,CCL11,CCL15,CCL17,CCL25 | 0.000447 | 0.16479 |
| sJIA-PAP vs all sJIA | GO_LYMPHOCYTE_CHEMOTAXIS | 64 | 34 | 5 | CCL7,CCL11,CCL15,CCL17,CCL25 | 0.00046 | 0.16479 |

**Table S6 Pathology and immunostaining characteristics of patient lung specimens**

| Patient Code | Diagnosis | Age Range (Years) | Major histologic findings | IL-18 | ICAM-5 | MMP-7 | CCL2 | Galectin-3 | CCL17 |
|---|---|---|---|---|---|---|---|---|---|
| 1 | sJIA | 0-5 | PAP/ELP | ++ | +++ | ++ | +++ | +++ | + |
| 2 | sJIA | 0-5 | PAP | ND | ++ | + | +++ | +++ | ND |
| 3 | sJIA | 11-15 | ELP | +++ | ND | + | ++ | + | ND |
| 4* | sJIA | 0-5 | PAP | ++ | + | + | ND | + | ND |
| 5 | sJIA | 0-5 | PAP/ELP | ND | ++ | + | ND | +++ | ND |
| 6 | sJIA | 0-5 | PAP | ND | + | ++ | ND | ND | - |
| 7 | sJIA | 0-5 | Limited ELP | ND | ++ | - | ND | ND | ND |
| 8 | sJIA | 6-10 | PAP/ELP | + | ++ | + | ND | ND | + |
| 8* | | 6-10 | PAP | + | ++ | + | | | + |
| 9 | *GATA2* | 16-20 | PAP/ELP/Fibrosis/PHTN | ++ | +++ | ++ | +++ | +++ | ND |
| 10 | *SFTPC* | 0-5 | Chronic pneumonitis of Infancy | ++ | ++ | + | ++ | +++ | ND |
| 11* | *NKX2.1* | 0-5 | ELP/Fibrosis | +++ | ND | ++ | ++ | ++ | ND |
| 12 | Idiopathic ILD | 6-10 | Chronic bronchiolitis/PAP | ++ | + | + | ++ | +++ | ND |
| 13 | Control | 11-15 | Asthma/Aspiration | + | ++ | -/+ | ++ | +++ | ND |
| 14* | Control | 0-5 | Acute bronchopneumonia | - | + | -/+ | +++ | +++ | ND |
| 15 | Control | 0-5 | Lung adjacent to tumor | + | + | + | ND | ND | + |

* Lung sample obtained at autopsy; remainder are surgical lung biopsies

PAP, pulmonary alveolar proteinosis; ELP, endogenous lipoid pneumonia; NSIP, non-specific interstitial pneumonia; VOD, veno-occlusive disease; PHTN, pulmonary hypertension; ND, not done due to limited material.

+, rare/few foci/mild; ++, occasional/scattered foci/moderate; +++, frequent/many foci/severe

**Table S7: Demographics for the validation cohort**

| Patient Group | sample type | Sample # | Age, median (IQR) | Female % |
|---|---|---|---|---|
| Ctrl (plasma) | plasma | 6 | NA* | NA* |
| Inactive sJIA | serum | 11 | 11 (5-14) | 85% (11/13) |
| | plasma | 4 | 10 (5-15) | 50% (2/4) |
| Active sJIA | serum | 10 | 7 (4.4-8.9) | 50% (5/10) |
| | plasma | 6 | 7.5 (4.5-12) | 33% (2/6) |
| MAS | serum | 9 | 5 (2.2-16) | 75% (6/8) |
| | plasma | 6 | 13 (10-16) | 83% (5/6) |
| sJIA-PAP** | serum | 13 | 7.6 (5.5-10) | 53% (8/15) |
| | plasma | 7 | 6 (2.8-10) | 43% (3/7) |
| Other LD | serum | 5 | 19 (1-20) | 100% (5/5) |
| | plasma | 1 | 2 (2-2) | 100% (1/1) |
| sJIA-LD resolved | serum | 1 | 16 (16-16) | 100% (1/1) |

* Data not available **The disease characteristics for each cases of sJIA-PAP

#: The five samples came from two patients. For samples within each patient, the sJIA activity varied (including inactive sJIA, active sJIA and MAS, see large variations in Figure 5A). However, their MMP7/ICAM5 levels remained

### Table S8: Protein abundance alterations in other types of lung disease identified in the literature

| | Serum | | | | BAL fluid | |
|---|---|---|---|---|---|---|
| **Gene Name** | **sJIA-PAP** | **IPF Study 1** | **IPF Study 2** | **RA-ILD*** | **NEHI** | **Surfactant mutant** |
| **CCL11** | Significant | Not Significant | Not Significant | NA | Not Significant | Not Significant |
| **GDF15** | Significant | Not Significant | Significant | NA | Not Significant | Not Significant |
| **CCL17** | Significant | Not Significant | Significant | Significant | Significant | Significant |
| **CCL25** | Significant | Not Significant | Not Significant | NA | Not Significant | Not Significant |
| **CCL7** | Significant | Not Significant | Not Significant | NA | Significant | Significant |
| **ICAM5** | Significant | Significant | Significant | Significant | Significant | Not Significant |
| **LGALS3** | Significant | Not Significant | Not Significant | Significant | Not Significant | Not Significant |
| **MMP7** | Significant | Significant | Not Significant | Significant | Not Significant | Significant |
| **Ref Number** | | 23 | 24 | 38 | 39 | 39 |

**"Significant" means:**

**1. The author identified the protein abundance change and thus reported the summary statistics**

**AND 2. the summary statistics pass the significance threshold as our study (sJIA-PAP): FDR < 20% and minimal fold change of 1.5.**

***: This study is only available thru an abstract, thus without full list of proteins with abundance change. We cannot determine whether other unreported proteins reached significance threshold in abundance change, so NA is reported.**
