## supplemental text for "Serum proteome analysis of systemic JIA and related pulmonary alveolar proteinosis identifies distinct inflammatory programs and biomarkers"

### Supplemental Methods

#### Cohort and disease definitions

Human subject protocols were approved by and conducted through the National Institutes of Allergy and Infectious Diseases (NIAID), Cincinnati Children's Hospital, Stanford University, the University of Pittsburgh, Seattle Children's Hospital, and Boston Children's Hospital. The patients represented 10 patient groups, based on clinical characterizations (**Table 1, Supplementary Table 1**). Classification as sJIA was based on a Childhood Arthritis and Rheumatology Research Alliance (CARRA)-initiated modification of the ILAR criteria[1] such that arthritis was not required, and MAS classification was by the 2016 classification criteria[2]. Active and inactive sJIA were defined by the submitting investigators. sJIA-PAP patients were known or strongly suspected to have the syndrome of digital clubbing and PAP-variant parenchymal lung disease observed in sJIA[3-6] and unresolved pulmonary disease at the time of sampling in the opinion of the submitting investigator. All sJIA-PAP patient samples were collected during treatment with IL-1 or IL-6 inhibitors (**Supplementary Table 1**). After running SOMAscan, the sJIA-PAP samples were divided into those with high levels of ferritin and CRP (sJIA-PAP$^{FCHi}$) and those without (sJIA-PAP$^{FCLo}$). To make this division, we set RFU thresholds for CRP and ferritin (**Supplementary Figures 3B-E**). CRP thresholds were selected based on values separating the top quintile of inactive sJIA from the bottom quintile of active sJIA. Ferritin thresholds were set similarly in the MAS group. NLRC4-MAS patients all carried known pathogenic mutations, and active disease was defined by the submitting investigator. NOMID (Neonatal-Onset Multisystem Inflammatory Disease) and SAVI (STING-Associated Vasculopathy of Infancy) samples were obtained prior to treatment with IL-1 blockade or JAK inhibition, respectively. All SAVI patients had interstitial lung disease. Primary PAP samples were derived from patients with autoimmune (anti-GM-CSF autoantibodies, n=10) and hereditary (surfactant protein deficiencies, n=4) causes for PAP. Healthy control samples derived from unaffected siblings and seronegative children from an unrelated asymptomatic *Pneumocystis* serology study[7].

The 5 main groups in the SOMAscan discovery cohort (healthy, inactive sJIA, active sJIA, MAS, sJIA-PAP) were represented in the validation cohort. In addition, the validation cohort included a group of "other lung disease" patients, who were known to have non-PAP lung disease at the time of sampling (e.g., idiopathic pulmonary hypertension or transient pneumonia). One sJIA patient with resolved lung disease was also identified in the validation cohort (**Supplementary Tables 1 and 7**).

#### Serum proteome analysis

We included 151 serum samples from 120 patients for proteomic profiling using SOMAscan. Most samples (78%) and patients (85%) were from five main groups (21 healthy controls, 28 inactive sJIA, 24 active sJIA, 10 MAS secondary to sJIA, and 10 sJIA-PAP). For samples from a patient belonging to different patient groups at different times (e.g., active MAS and inactive sJIA, sJIA-PAP$^{FCHi}$ and sJIA-PAP$^{FCLo}$), the samples were assigned to the separate patient groups. Serum samples were distributed by disease group equally between two plates and measured by SOMAscan assay, according to the manufacturer's instructions, (SOMAlogic, Boulder, CO) in collaboration with the NIH Center for Human Immunology (CHI). SOMAscan data were first normalized to remove hybridization variation within the run, followed by median normalization across all samples to remove other assay biases within the run and finally calibrated to remove assay differences between runs. 187 of 1317 targets were flagged for between-plate calibrator variation but remained in the analysis. Flagged SOMAmers were not enriched in any differentially expressed target lists. Post-normalization, all samples met predefined acceptance criteria and were included in the analysis. A complete description of the normalization and calibration procedures is available at: www.somalogic.com/wp-content/uploads/2017/03/SSM-071-Rev-0-Technical-Note-SOMAscan-Data-Standardization.pdf. Quantitative assessment of target abundance was expressed as relative fluorescence units (RFU). IL-18, IL-18 binding protein, and CXCL9 were measured by Luminex assay as described previously[8] and their values were also expressed as RFU. As IL-18 was not included as part of SOMAscan panel, we combined the RFU values for IL-18 from the Luminex assay with the RFU data matrix generated by SOMAscan. The p-values for IL-18 results and SOMAscan results were adjusted for multiple tests. Leftover serum from 80 samples remaining after SOMAscan analysis were re-analyzed for ICAM5 by ELISA per manufacturer's instructions (R&D Systems). Additionally, a validation cohort (49 serum samples and 29 plasma samples) was assayed for IL-18 and CXCL9 by Luminex and for ICAM5 and MMP7 by ELISA (R&D systems).

**Mapping SOMAscan probes to gene symbol IDs**

For 45 analytes, a SOMAscan probe matched to a protein complex. In these cases, we chose one protein from the complex to represent the probe (**Supplementary Tables 2-4)**. If multiple probes matched to the same gene symbol, their values were averaged for down-stream analyses.

**Linear regression-based modeling analysis**

The experimental and analytical scheme of the whole study is summarized in **Supplementary Figure 2**.

Some patients contributed multiple samples to the discovery cohort, although no sJIA-PAP samples were collected at onset of sJIA or PAP. For LIMMA (Linear Models for Microarray Data) analysis, to represent the variations between disease groups, we pooled all results belonging to the same individual within a patient group and averaged their protein abundance.

While most patients were in early childhood (all main patient groups' median age was <10 years old, **Table 1**), a few were adults (including one sJIA-PAP patient). The small number of adult patients may not capture the full variety in the adult diseases. Thus, we focused on the pediatric population in the LIMMA model and included only patients with age <18 years old, accounting for 86% patients from the 5 main patient groups (see above). Adding adult patients back into the comparisons did not change interpretation, and all boxplots (e.g., **Supplementary Figure 10** and violin plots (e.g., **Figure 1D**) include adult patients.

| **Subjects removed from the LIMMA analysis due to age higher than 18** | |
|---|---|
| **Groups** | **Removed** |
| Healthy Ctrl | 4/21 |
| Inactive sJIA | 4/28 |
| Active sJIA | 2/24 |
| Active MAS | 2/10 |
| sJIA-PAP | |
| subgroup: sJIA-PAP FCLow | 1/8 |
| subgroup: sJIA-PAP FCHigh | 1/6 |

Patients in groups with MAS or sJIA-PAP had multiple disease components: for example, some patients with sJIA-PAP also had an active sJIA disease component. We distinguished patient groups from disease components. Note that disease and component names may overlap (e.g., the sJIA group vs. the sJIA disease component) and disease components can appear in other patient groups (e.g., MAS disease component in sJIA-PAP patient samples). To capture the serum proteome alterations attributed to a specific disease component (sJIA, MAS, or sJIA-PAP), we used LIMMA, in which the overall proteome alteration was regressed against the disease component(s) present in each individual patient (**Supplementary Figures 2A-B**). A model matrix was used to represent the disease component constitution for each patient.

When generating disease components for inactive sJIA, active sJIA and MAS, we made comparisons between controls, inactive sJIA, active sJIA, and MAS, which were well-characterized according to classification criteria, as described above. The disease components in these samples were represented by categorical values (0 or 1) in the model matrix.

We generated the sJIA-PAP disease component differently, as this component was more complex. Classical MAS activity markers, represented in the MAS serum activity score, were variable in the sJIA-PAP patients (**Supplementary Figures 3C-E and Figure 1D**). Thus, using a categorical classification (0 or 1) of MAS activity in the sJIA-PAP patient group would lead to under- or over-estimation of MAS activity. To account for this, we treated MAS activity as a continuous variable (the MAS serum activity score) in the LIMMA model when generating the sJIA-PAP disease activity component (**Supplementary Figure 2C**).

Potential confounders (sex, age, and visual hemolysis using 0-4 ordinal scale) were included in generating the component scores. Twelve of 151 samples showed signs of hemolysis (score >0). Standard LIMMA procedure was implemented[9] using Bayesian statistics. Significance thresholds for this discovery analysis were set at 1.5-fold absolute differences and Benjamin-Hochberg FDR ≤20%. A substantial fraction of targets met a more stringent FDR threshold (**Supplementary Tables 2-4;** Adj. P value=FDR; $Log_2FC$=Coefficients of concentration change from LIMMA analysis).

We used R to perform all statistical analysis. LIMMA analysis was performed by using limma package from Bioconductor[10]. The LIMMA analysis produces its own P estimation. The p-values for all between-group comparisons, and for correlations of component scores, were calculated using ranking based statistics to avoid assumption of normal distribution, as described in the figure legends.

**The disease component specific serum activity score**

We calculated a score using significantly different proteins associated with a disease component to summarize the serum proteome profile for each sample. For a given serum sample and a given disease component, we calculated the sum of log2 abundance (effectively the log2 of the product) for the over-expressed proteins and under-expressed proteins separately. The differences between the sums were divided by the number of proteins used. The equation is very similar to the one for calculating the geometric mean of all over-expressed and under-expressed proteins, except that the under-expressed proteins were given a negative sign to reflect the fact that their expression levels are reduced by the disease component.

$$S_i = \frac{log2[\prod_{Prot \, \epsilon \, pos} x_i(prot.)] - log2[\prod_{Prot \, \in \, neg} x_i(prot.)]}{N_{pos} + N_{neg}}$$

i, sample id.
Xi, abundance of a given protein in sample i.
N, protein number.
Pos/neg, significantly over or under-expressed proteins associated with a given disease component.

**Longitudinal analysis**

Serum activity scores or levels of individual proteins were analyzed in sJIA-PAP patients with longitudinal samples. For these individual samples, to evaluate which values significantly deviated from the base condition (active sJIA), we pooled the samples in the active sJIA group (without sJIA-PAP or MAS) to derive a distribution representing the population variations within the base condition. Z scores (number of standard deviations from the mean of the base condition) were calculated, with Z=1.96 as the upper limit of the confidence interval for the given value in the active sJIA population.

**Functional annotation of the proteins**

We tested the over-presentation of 7573 gene sets from Gene Ontology Biological Processes in the gene lists (**Supplementary Tables 2-4**) using a hypergeometric test, with all proteins measured as the background. The minimum size for the gene sets to be tested was 5. Adjusted p-values were calculated using the Benjamini-Hochberg false discovery rate (FDR) method. Results of this analysis are presented in **Supplementary Table 5.**

We also used the following GO terms to find cytokines and chemokines: GO:0005125: cytokine activity; GO:0042056: chemoattractant activity. Gene symbol IDs were used for GO term matches.

**Transcriptional data from GTEx**

Organ-specific transcriptional data presented in **Figure 4A** were retrieved from the Genome-Tissue Expression (GTex) data portal[11] and ranked by transcript abundance in the organs.

**Immunofluorescence and immunohistochemistry**

Patient lung samples for the study were from archived material performed for clinical diagnosis and approved for use in this study by Seattle Children's Hospital Institutional Review Board. Pathologic interpretation of seven of eight sJIA samples (**Supplemental Table 6**, patient code 2-8) was performed by the originating institution with re-review by three experienced pediatric pulmonary pathologists to issue a primary diagnosis. The primary pathologist (GD) has over 20 years' experience interpreting pediatric lung pathology. The remaining cases (patient codes 1, 9-15) originated from Seattle Children's Hospital and were identified from a pathology database as "ILD" (genetic disorders in surfactant metabolism and idiopathic ILD) and "controls". The clinical pathology interpretation from these cases was re-reviewed by GD to arrive at a primary diagnosis; in no instances was the original diagnosis changed.

Immunohistochemical staining was carried out on adjacent 5 micron sections from paraffin-embedded tissue. MMP7, Galectin-3 and CCL2 were performed on the Ventana BenchMark ULTRA immunostainer (Roche) after antigen retrieval with citrate buffer, pH 6.0 and endogenous biotin block.

IL-18, ICAM5 and CCL17 antibodies were incubated overnight at room temperature, followed by biotinylated antibody (1:500) and streptavidin-biotin-peroxidase complex, and peroxidase activity was visualized using diaminobenzidine (all Vector Laboratories). Negative controls consisted of sections incubated without primary antibody. ICAM5 and MMP7 were co-stained with cytokeratin (CK) 7 or CD163 by immunofluorescence; ICAM5 and MMP7 were developed with donkey anti-rabbit IgG, fluorescein conjugate and cytokeratin 7 and CD163 by donkey anti-mouse IgG, cyanine (Cy3) conjugate (all 1:500, Jackson ImmunoResearch Laboratories). Coverslips were mounted using Vectashield fluorescent mounting medium with DAPI nuclear stain (Vector Laboratories). Images were captured with a digital camera mounted on a Nikon Eclipse 80i microscope and using NIS-Elements Advanced Research Software v4.13 (Nikon Instruments Inc.).

There was no formal scoring system of specific pathologic findings. Interpretation of the immunohistochemistry was performed blinded to the diagnosis with the following scoring system: +, rare/few foci/mild; ++, occasional/scattered foci/moderate; +++, frequent/many foci/severe.

**Staining Reagents**

| Target | Species | Dilution | Company (Catalog #) |
|---|---|---|---|
| IL-18 | Rabbit | 1:250 | EMD Millipore (06-115) |
| ICAM5 | Rabbit polyclonal | 1:100 | Sigma-Aldrich (HPA009083) |
| MMP7 | Rabbit polyclonal | 1:500 | GeneTex (GTX32725) |
| CCL2 | Mouse monoclonal | 1:1000 | Novus biologicals (NBP2-22115) |
| Galectin-3 | Mouse monoclonal | 1:1200 | Novus biologicals (NBP-92690) |
| CCL17/TARC | Goat polyclonal | 1:500 | R&D Systems (AF364) |
| Cytokeratin 7 | Mouse monoclonal | 1:100 | Leica Biosystems (CK7-560-L-CE) |
| CD163 | Mouse monoclonal | 1:100 | Leica Biosystems (CD163-L-CE) |

**Analysis of cell-type specific RNA levels in normal adult lung**

The processed 10x sequencing data of the human lung cell atlas presented in **Supplementary Figure 15** was downloaded from Synapse, accession syn21041850[12], and analyzed using Seurat version 3.1.5.

**Data availability**

The SOMAscan data will be uploaded to GEO (Gene Expression Omnibus) upon publication.

**Statistical Analysis**

Details for linear regression analysis by LIMMA are described above. Other statistical analyses were performed using ranking-based approaches (such as Wilcoxon test or Spearman's correlation). Details for each analysis are provided in the relevant Figure legend.

**Ethics approval**

All activities were performed in compliance with the Helsinki Declaration under protocols approved by institutional review boards at the following institutions: Stanford University, University of Cincinnati, University of Pittsburgh, National Institute of Allergy and Infectious

Diseases at the National Institutes of Health, Seattle Children's Hospital, and Boston Children's Hospital. All subjects, or their legal guardians, gave informed consent.
